# Bibliometric Insights into Thirty-Nine (39) Years of Research on *Pseudomonas aeruginosa* in Diabetic Foot Infections

**DOI:** 10.1101/2025.10.14.25337998

**Authors:** Morumda Daji, Dauda Wadzani Palnam, Peter Abraham, Israel Ogwuche Ogra, Dogara Elisha Tumba, Emohchonne Utos Jonathan, Elkanah Glen, Okechukwu Kalu Iroha, Mercy Nathaniel, Samson Usman, Seun Cecilia Joshua, Mela Ilu Luka, Grace Peter Wabba, Naanswan Joseph Dasoem, Ndukwe K. Johnson, Emmanuel Oluwadare Balogun, Zainab Kasim Mohammed, Umezuruike Linus Opara

## Abstract

The ability of *Pseudomonas aeruginosa* to form biofilms and develop multidrug resistance has made it a highly significant pathogen in diabetic foot infections (DFIs), contributing to global morbidity. This research carried out a systematic review and bibliometric analysis of 873 articles on *P. aeruginosa* in DFIs, published between 1985 and 2024, that were retrieved from the Scopus database in order to clarify changing research trends and pinpoint areas of innovation. Bibliometric tools including Bibliometrix in R and VOSviewer were used to map authorship networks, keyword co-occurrence, and thematic evolution. After language and document-type filtering and subsequent manual keyword screening, 333 relevant publications were analyzed. Research output has significantly increased over the past two decades with an annual growth rate of 9.85%, and a noticeable shift in focus from general infection studies to focused studies on molecular mechanisms, antibiotic resistance, and sophisticated therapeutic interventions. Innovation hotspots have been identified in biofilm research, antimicrobial resistance, and new treatments like phage therapy and quorum quenching. Global research distribution is still unequal despite these advancements with most publications coming from nations with strong research infrastructure, while contributions from less resource-rich areas are scarce. Additionally, collaboration patterns show fragmented international cooperation and a preference for intra-country partnerships. Emerging innovation hotspots include anti-biofilm strategies, genomics-driven resistance profiling, and next-generation therapeutic development. However, the study observes systemic disparities in research equity and knowledge sharing. Addressing these issues through inclusive research partnerships and stronger policy-to-practice integration is essential to advancing effective DFI management globally. To make future advancements in the control of *P. aeruginosa* in diabetic foot infections, it will be important to address these disparities through inclusive and cross-disciplinary research.

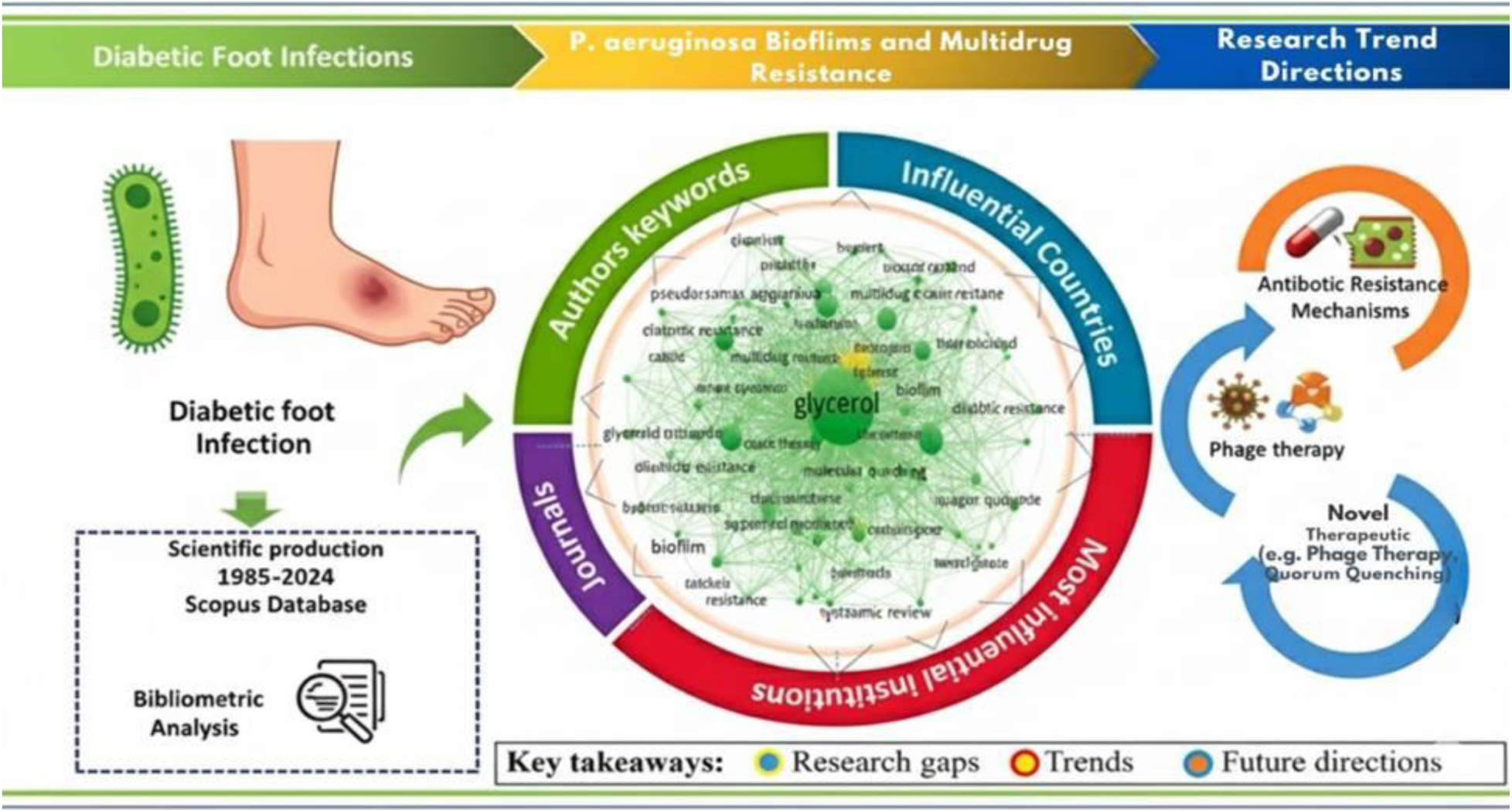

## Introduction

Out of the approximately 537 million individuals across the globe who are affected by diabetes, a range of 19% to 34% will experience the development of a diabetic foot ulcer (DFU) at some point in their lives. Around 20% of individuals who experience a diabetic foot ulcer (DFU) may need to undergo amputation of the lower extremity, either as a minor procedure below the ankle or a major procedure above the ankle, or both. Additionally, 10% of these individuals will pass away within one year of being diagnosed with their initial DFU (McDermott *et al*., 2023). Diabetic foot infections (DFIs) are a significant and troublesome consequence of diabetes mellitus, linked to high rates of illness and the potential for amputations due to the wide range of microorganisms involved and their resistance patterns (Mansour and Abdulwaha, 2024).

*Pseudomonas aeruginosa* is a leading cause of opportunistic infections in humans, causing a variety of acute infections. This bacterium has a versatile metabolic capability that enables it to outcompete other bacteria in the nutrient-limited environment of the hospital, thereby making it the primary cause of nosocomial infection-related morbidity and mortality (Lee *et al.,* 2023). *P. aeruginosa* is a member of the "ESKAPE" pathogens, which include *Enterobacter* species, *Staphlylococcus aureus, Klebsiella pneumoniae, Acinetobacter baumannii*, and *Enterococcus fecium* which are clinically concerning because of their broad-spectrum antibiotic resistance profiles and potential to cause fatal nosocomial infections (De Oliveira *et al.,* 2020). Additionally, P. aeruginosa has been placed in the "critical" tier (priority 1 group among critical, high, and medium priorities) on the WHO priority list of antibiotic-resistant bacteria requiring research and development of effective treatments. *P. aeruginosa’s* high and broad resistance profiles, which include MDR and XDR, were ascribed to the interaction of acquired, adaptive, and innate resistance mechanisms (Lee *et al.,* 2023). *P. aeruginosa* is a major contributor to all diabetes-associated sepsis and amputation in this patient population, as it causes substantial harm to individuals with diabetic foot ulcers. *P. aeruginosa* is more commonly seen in chronic wound infections than other microbes, where it slows wound healing by creating biofilm in the wound, which makes it stronger and more difficult to heal (Garousi *et al.,* 2023). Also, biofilm production together with the release of other virulence factors such as catalases, rhamnolipids, lipases, proteases, pyocyanin, pyoverdine, and exopolysaccharides, with the majority of them governed by quorum-sensing (QS) systems, which include Las, Rhl, QscR, Pqs, and Iqs, has empowered *P. aeruginosa* to successfully infect the cells of the host and also evade the immune system of the host, strengthening its pathogenicity (Chichón *et al.,* 2023).

However, the large amount of data on diabetic foot brought about by the quick increase in publication output appears to be making it harder to understand the new patterns and areas of study interest. Conventional reviews, on the other hand, are limited in fully and methodically presenting the topic’s knowledge mapping. A new method for examining the published scientific literature is bibliometric analysis (Zhao *et al.,* 2023). Although knowledge mapping studies on diabetic foot have been done (Zhao *et al.,* 2023), they were based on the Web of Science database and did not capture diabetic foot infection and were unable to involve *Pseudomonas aeruginosa* precisely, even though it has a great impact on diabetic foot infection.

This paper provides an overview of global research output on *Pseudomonas aeruginosa* in diabetic foot infections, tracking its development, significant contributors, and areas of research interest with an intention to track the focus of intellectual energy, the key authors and what paradigms are emerging. In order to visualize collaboration networks and knowledge clusters, identify influential authors, institutions, and nations propelling the field forward, track the emergence of new topics and trends, and assess the impact of *Pseudomonas aeruginosa* on diabetic foot infection, we used network analysis, citation metrics, and thematic analysis on a large dataset of scientific publications. To attain the stated objective of this study, the following research questions were used to lead the study:

RQ1: How has global research on *P. aeruginosa* evolved in focus and intensity over the last two decades?
RQ2: Which subdomains (e.g., biofilm research, antimicrobial resistance, phage therapy) are emerging as hotbeds of innovation?
RQ3: How equitable is the global distribution of *P. aeruginosa* research, and what are the patterns of collaboration?

## Methodology

This work aims to address a gap in the literature by combining quantitative (bibliometric) and qualitative (systematic review) analyses to elucidate trends in research on Pseudomonas aeruginosa in diabetic foot infections. The technique outlined in this work is illustrated in Fig. 1, which delineates the roadmap for the systematic review of literature and the conducted bibliometrics. The process commences with the criteria for the inclusion or exclusion of studies, followed by the selection of scientific journal databases, pertinent keywords, and search algorithms. This is succeeded by the Systematic Literature Review (SLR) adhering to the PRISMA 2020 guidelines, culminating in the analysis of the results (Bibliometrics) utilizing software tools. Evaluate Outcomes Utilizing Scopus and Bibliometrix via R.

**Fig 1A:**
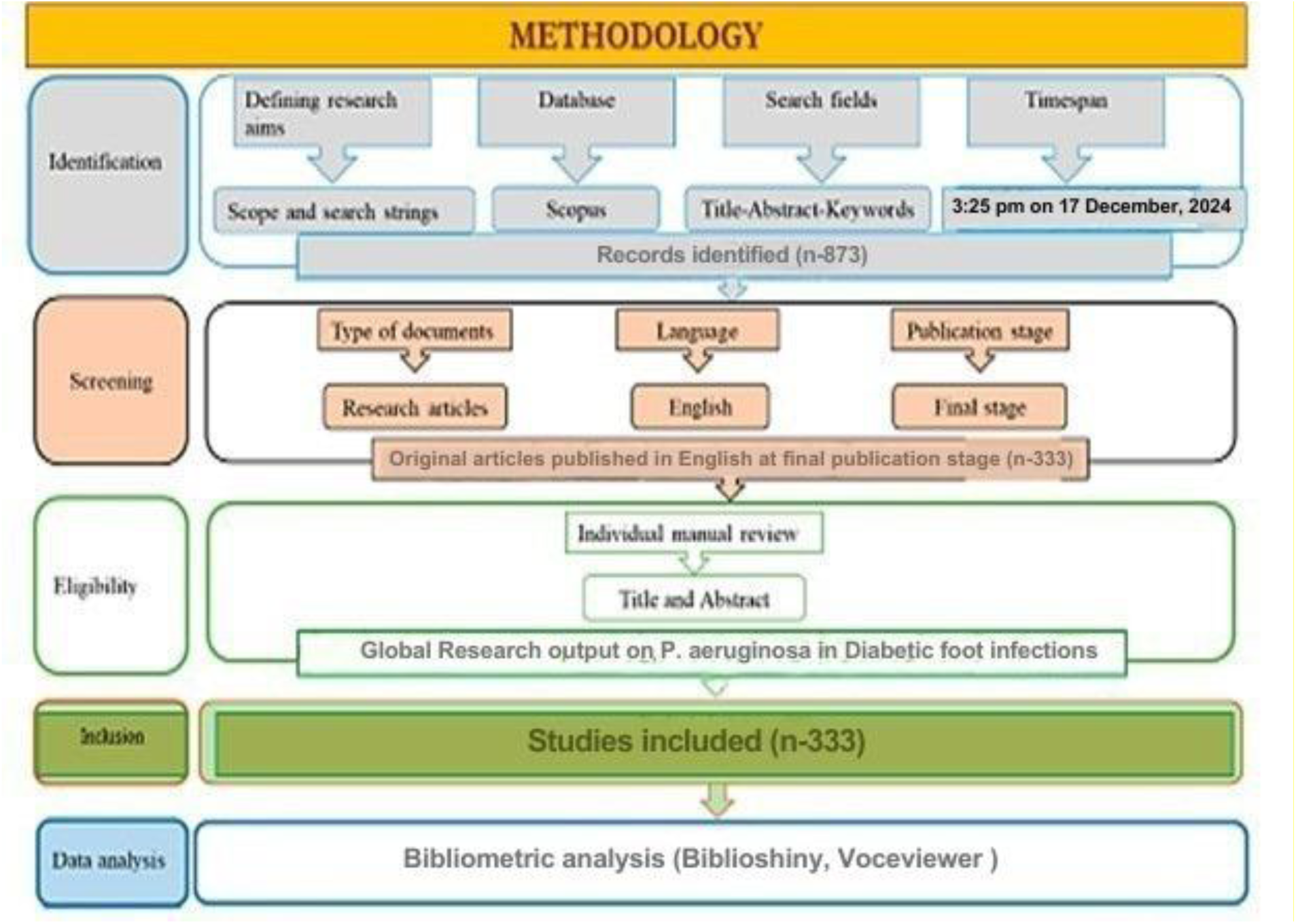
Conceptual framework of Global research output on *Pseudomonas aeruginosa* in diabetic foot infections

### 2.1. Search of the Literature

We performed an exhaustive literature search in the Scopus database to investigate global publications about Pseudomonas aeruginosa in diabetic foot infections, concentrating on studies published from 1985 to December 17, 2024, at around 3:25 PM. The search approach included targeted search strings within the "Title-Abstract-Keywords" fields. These strings incorporated a combination of Boolean operators (OR and AND), truncation symbols (*), and quote marks (“. . .”) to guarantee thorough and pertinent retrieval. We employed the subsequent search strings: ("Pseudomonas aeruginosa" OR "P. aeruginosa") AND ("diabetic foot" OR "diabetic feet" OR "diabetic foot infection" OR "DFI" OR "diabetic foot ulcer" OR "DFU" OR "foot ulcer" OR "foot infection") AND (PUBYEAR >= 1985 AND PUBYEAR <= 2024).

### 2.2. Study selection

A systematic literature review (SLR) was done to enable the selection of relevant works according to the flow diagram of the PRISMA 2020 declaration, which indicates the number of studies identified, eliminated for duplicates, reviewed for eligibility, and eventually included in the revision. The selection of literature for this review is premised on a careful study of the titles and abstracts acquired from the initial search results to determine their alignment with the stated inclusion criteria. Those entries that match these requirements are further reviewed by a careful study of the full texts when appropriate to confirm their eligibility. This systematic procedure does not involve any software or artificial intelligence (AI) techniques; rather, it depends only on manual analysis and expert judgment to assure the integrity and quality of the literature selected for inclusion in the review.

### 2.3 Inclusion Criteria

Publications that were directly related to the study focus on *Pseudomonas aeruginosa* in diabetic foot infections (DFIs) were included in this bibliometric review. Documents that specifically discussed *P. aeruginosa*’s role, pathogenic mechanisms, epidemiology, antimicrobial resistance, or treatment approaches in relation to diabetic foot ulcers or infections were chosen. Original research articles, review papers, conference proceedings, and editorials were all included as long as they underwent peer review and offered significant scientific contributions to the field. To ensure consistency in analysis and interpretation, only English-language publications were taken into consideration. To further capture the historical development, thematic evolution, and current advancements in the field, the inclusion period was extended from 1985 to 2024. The Scopus database, which was selected for its thorough indexing of reputable, peer-reviewed journals in the biomedical and health sciences, provided the publications. The records that were chosen were those that contained pertinent keywords in the title, abstract, or keyword sections, such as "*Pseudomonas aeruginosa*," "diabetic foot," "DFI," and "diabetic foot ulcer." This ensured that the dataset for the bibliometric analysis was targeted and topic-specific. A CSV file format was then used to export the resultant records.

**Figure 2.1:** PRISMA model-guided flow chart of the search methodology

### 2.2. Analytical tools and preprocessing

The Bibliometrix package (v. 4.1.3) in R Studio software for thorough bibliometric analysis and VOSviewer (v. 1.6.19), a scientific knowledge mapping tool, were used to further analyze the retrieved dataset after it was exported in CSV format (Van Eck and Waltman, 2010; Aria and Cuccurullo, 2017; Sganzerla et al., 2021). Given the limits of these technologies in data preparation (Moral-Munoz et al., 2020), Microsoft Excel was applied for data cleaning to assure the dataset’s consistency, dependability, and correctness. Thereafter, a human check was undertaken to assure the database’s quality for further analysis.

### 2.3. Bibliometric analysis and visualization

Bibliometric analysis and mapping were conducted using VOSviewer and the Bibliometrix R package in RStudio on the screened dataset to identify dominant research topics, patterns of drug resistance, virulence variables, and geographic predominance. These findings guided the refining of search keywords, eligibility criteria, and research selection by emphasizing relevant topics and underexplored regions. To address RQ1, the following bibliometric techniques were used: Productivity and impact indicators (number of documents, citations, h-index, and m-index) were used to analyze scientific production on Pseudomonas aeruginosa in diabetic foot infections, with a focus on important contributors (authors, institutions or affiliations, and countries) (Gonzales-Malca et al., 2022). Additionally, scientific mapping analysis (Baker et al., 2021) was used to examine the relationships between these contributions, particularly the writers, and network visualization was used to further this investigation. Furthermore, thematic evolution analysis and keyword frequency and co-occurrence improved the identification and completion of research gaps and future directions in response to RQ2 and RQ3. By creating visual maps based on important keywords, authors, and their relationships, keyword co-occurrence network analysis was carried out to obtain a thorough grasp of the current study area. Additionally, using the methodology outlined by Cobo et al. (2011), maps were created to highlight scientific trends, study themes, and different dataset architectures.

## 3.0 Results

### 3.1 Comparison of results obtained from various databases

Table 1 shows the difference in the number of documents retrieved from databases on *Pseudomonas aeruginosa* in diabetic foot infection, such as Google Scholar, PubMed, the Lens, Dimensions, ProQuest, Web of Science, and Scopus. This gives vital insights into their extent, coverage, and relevance to the study topic. The highest number of documents on the research subject were retrieved from Dimensions (7,071), followed by ProQuest (6,851). The least were obtained from Google Scholar (167) and The Lens (5).

**Table 1:** Documents retrieved from databases on *Pseudomonas aeruginosa* in diabetic foot infection.

| Database | No of articles |
| --- | --- |
| Google scholar | 162 |
| PubMed | 349 |
| Lens | 5 |
| Dimensions | 7,071 |
| ProQuest | 6,851 |
| Scopus | 873 |
| Web of Science | 456 |

The Dimensions database has more materials on *Pseudomonas aeruginosa* in diabetic foot infections than other databases because it covers a larger spectrum of research outputs, including peer-reviewed publications, preprints, clinical trials, patents, and grants. Dimensions uses a broader indexing approach that covers both global and regional articles. It also incorporates material from open-access sites and preprint servers like bioRxiv and medRxiv, providing wider coverage of recent and developing research. Additionally, Dimensions leverages AI-driven semantic search and a linked data architecture, which facilitates the finding of related papers across many disciplines. This broad and adaptable methodology makes Dimensions particularly efficient for accessing a greater number of research on specific issues like P. aeruginosa in diabetic foot infections. ProQuest also has more because it brings together a huge number of dissertations, theses, conference papers, and full-text academic journals, many of which are from schools that aren’t listed in other databases. It has an advantage in finding a lot of topic-specific documents because it focuses on both academic and "grey" literature. Google Scholar is a big search engine, but it often only shows highly cited or core scholarly articles. Its results can also be limited by the availability of documents, the accuracy of metadata, and regional access controls. The Lens is mostly a database for patents and literature about new ideas, so it doesn’t find as many relevant scientific or clinical research articles, which is why its number is so low.

### 3.2 An Overview of Research on *Pseudomonas aeruginosa* in diabetic foot infections

Research Performance Statistics on Global research output on *Pseudomonas aeruginosa* in diabetic foot infections, as shown in Figure 1, revealed 333 publications from 1985 to 2024 with a 9.85% annual growth rate. This growth rate reveals a great interest and involvement of this pathogen in diabetic foot infection, as many researchers have attempted to study this disturbing condition. Also, this analysis revealed that only 8 publications are from single authors, while the majority (325) are from multiple authors. This reveals the growing interest and multifaceted approach aimed at tackling *Pseudomonas aeruginosa* in diabetic foot infection. The international coauthor rate of 15.62% shows that this co-authorship is regional, thereby demonstrating a need for increased international collaboration and networking so as to have a robust approach in developing an effective treatment option.

The distribution of document types on global research output on *Pseudomonas aeruginosa* in diabetic foot infections is shown in Figure 1B. Articles account for 92% (307) of the documents, while only 8% (26) are review articles. The bulk of publications are journal papers, which are frequently regarded as the gold standard of scientific communication. This shows a robust foundation of empirical and experimental research being undertaken and communicated. Review papers are the second and last publications, showcasing attempts to integrate current knowledge and map the state of the subject, a vital phase in a burgeoning research topic.

**Figure 1B:**
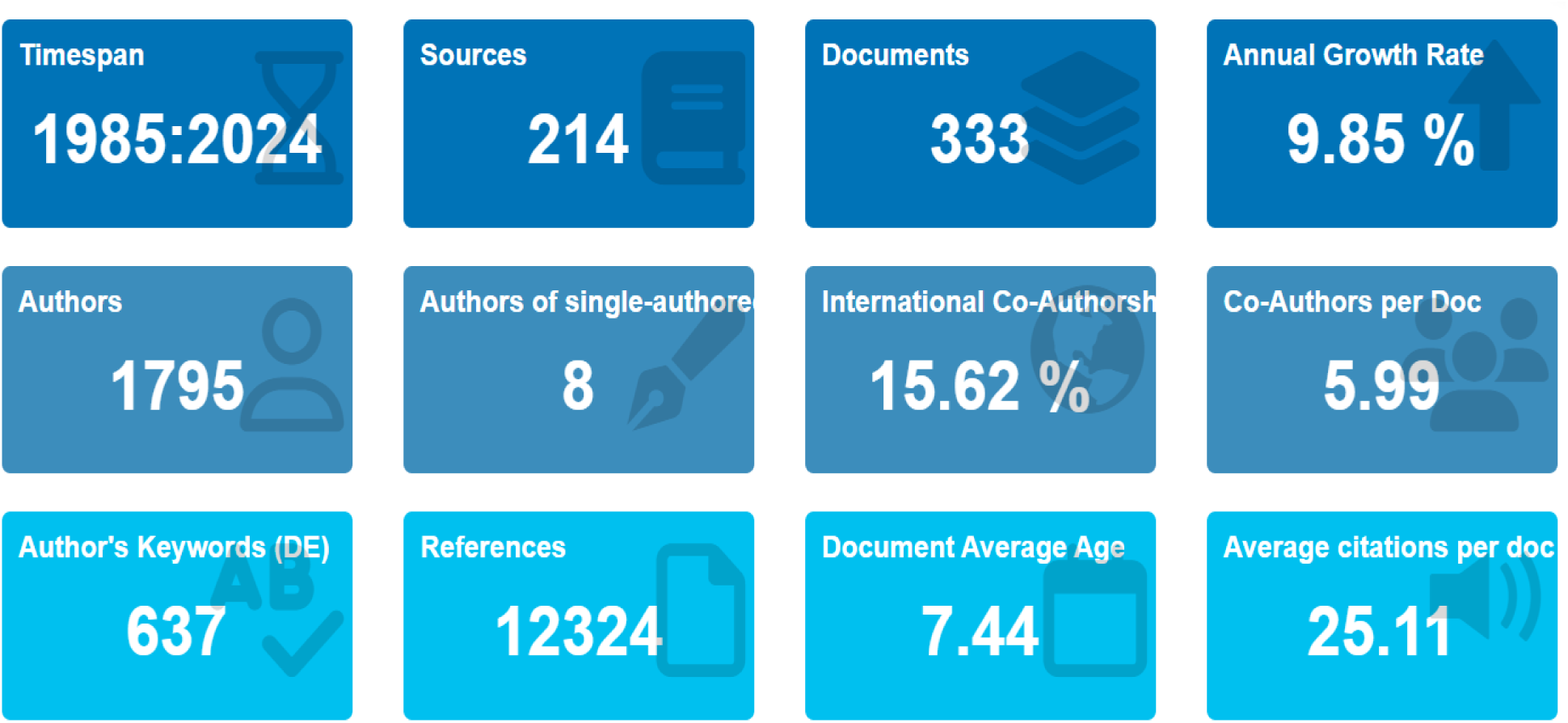
Main Information on Research Performance Statistics on Global Research Output on *Pseudomonas aeruginosa* in Diabetic Foot Infections.

**Figure 1C:**
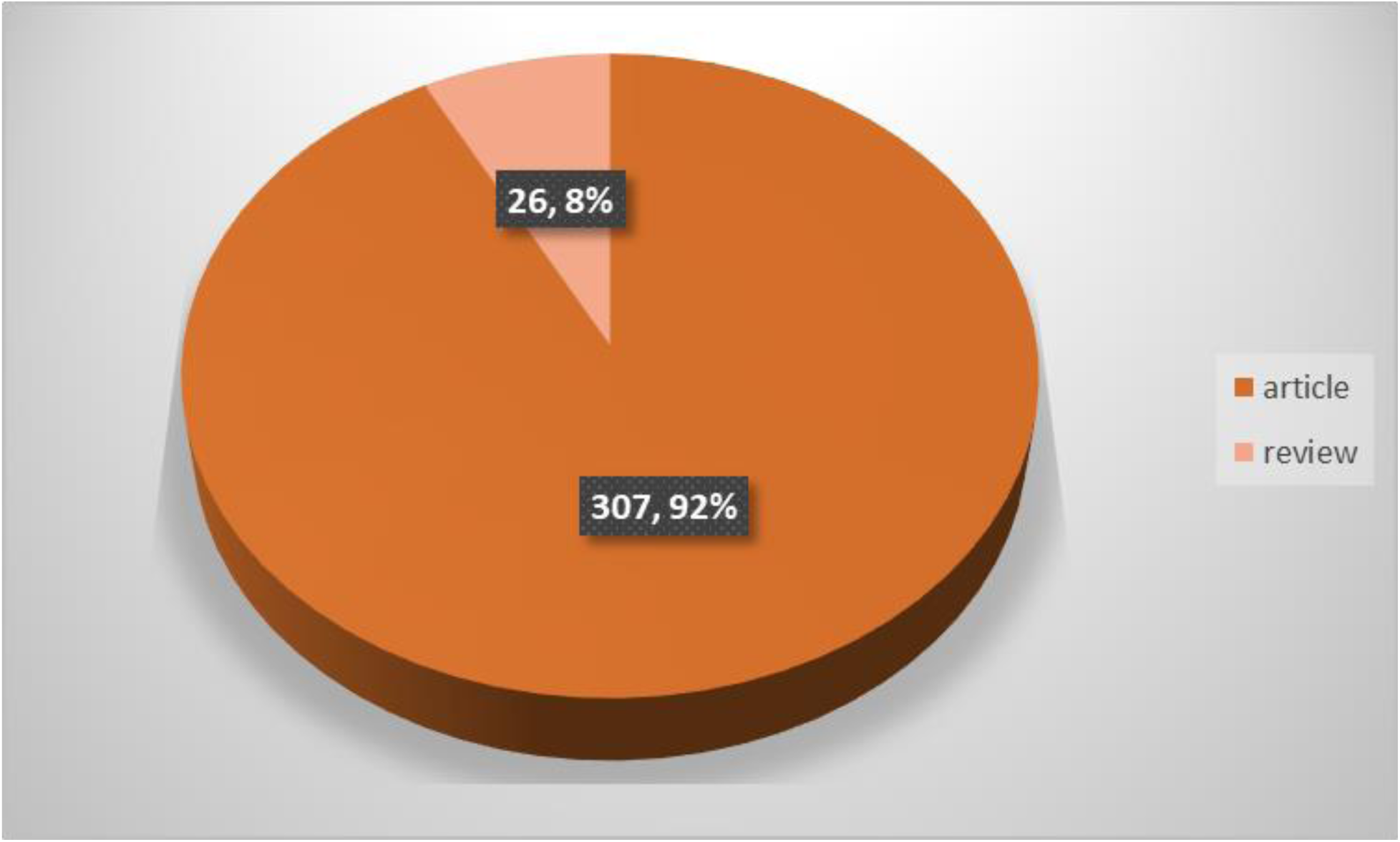
Distribution of document types on global research output on *Pseudomonas aeruginosa* in diabetic foot infections.

### 3.3 Publication trends on Global research output on *Pseudomonas aeruginosa* in diabetic foot infections

#### 3.3.1 Research publication trends

Figure 2 shows the trajectory of evolution of research trends on *Pseudomonas aeruginosa* in diabetic foot infections worldwide. Since 1985, the pathogen associated with this clinical state has been the focus of three major stages of increase in research output.

**Figure 2A:**
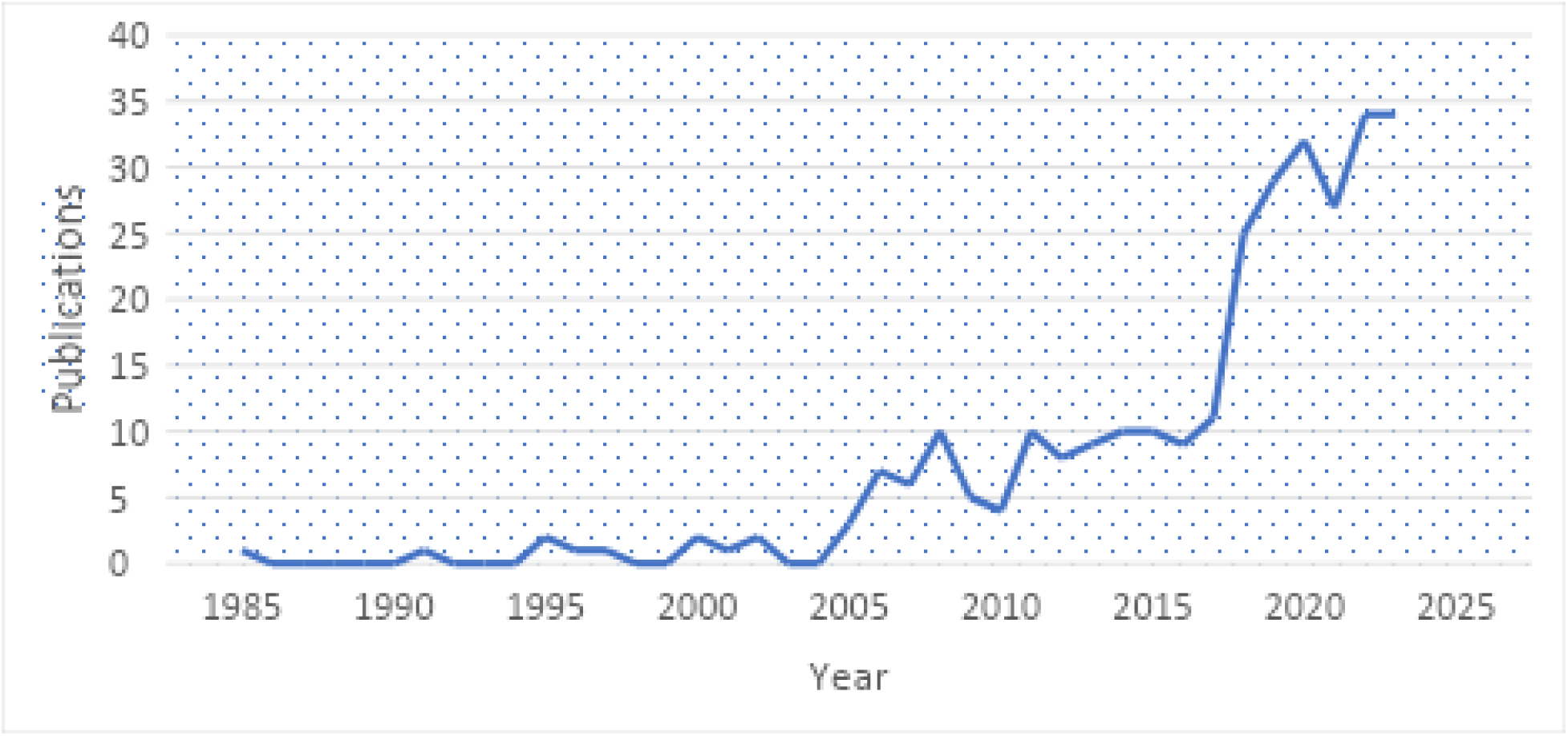
Annual Publication Production on *Pseudomonas aeruginosa* in diabetic foot infections

**Figure 2B:**
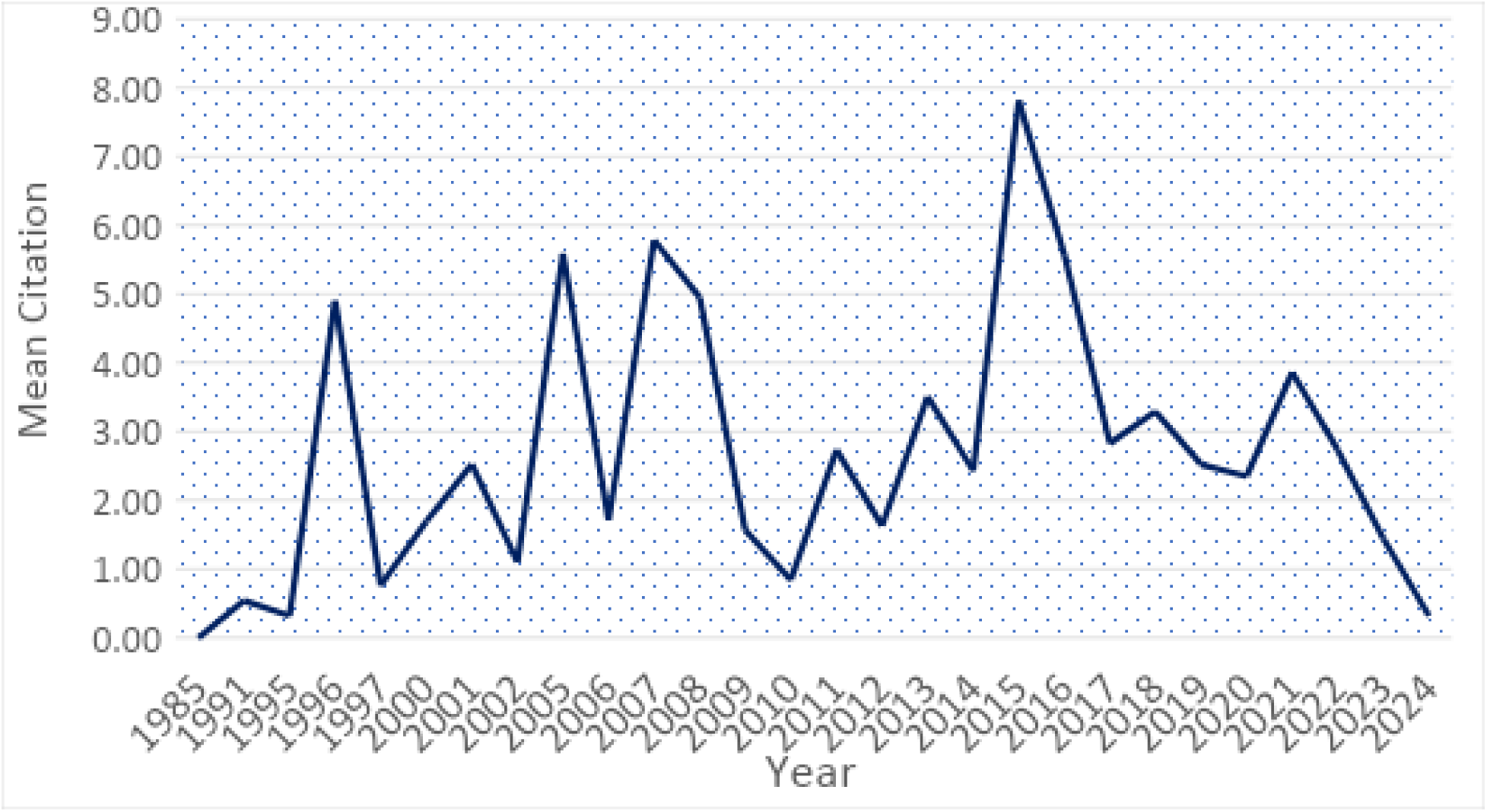
Yearly Mean Citations of Scientific Publications on *Pseudomonas aeruginosa* in diabetic foot infections

**Figure 2C:**
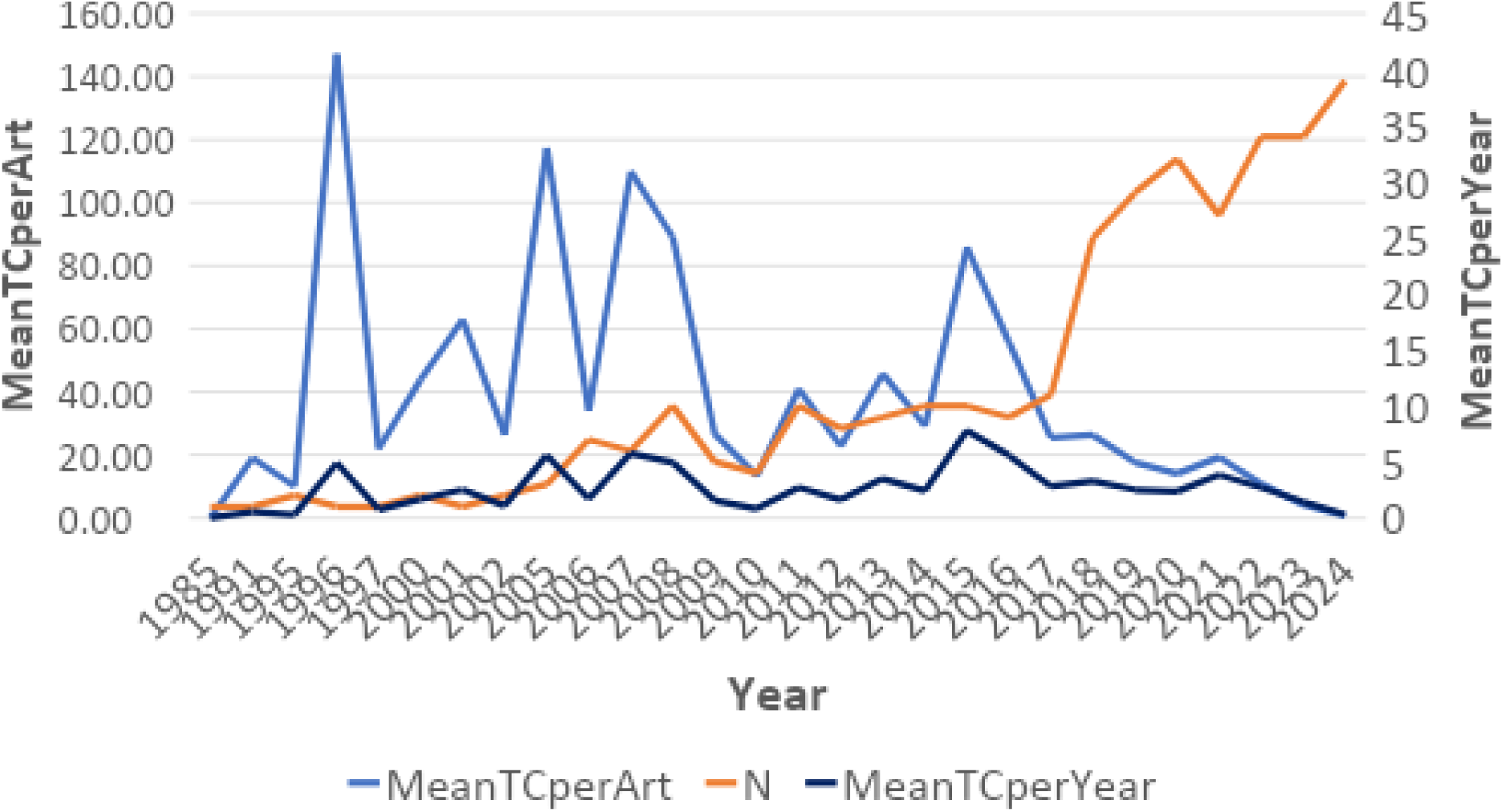
Annual Total Citation Per Year on *Pseudomonas aeruginosa* in diabetic foot infections

##### Early Stage (1985–2004): Foundation and Low Output

In the early phase, research on P. aeruginosa was scant and mainly vague, sometimes incorporated inside broader investigations on diabetic foot infections or general infectious disorders. The first article emerged in 1985, concentrating on antibacterial action, but it wasn’t until the early 1990s that interest slightly developed, albeit inconsistently. During this time, P. aeruginosa was not yet established as a significant concern in diabetic foot infections, and few studies particularly addressed it. The low number of publications and years of inactivity (e.g., 2003–2004) imply limited global acknowledgment of its therapeutic value in DFIs.

##### Growth Phase (2005–2020): Recognition of Clinical Relevance

This phase marks a large surge in research production, spurred by the rising awareness of *P. aeruginosa* as a common and problematic infection in diabetic foot ulcers. Publications continually grew, culminating in 2020 with 32 research. Research began concentrating on the polymicrobial nature of DFIs, *P. aeruginosa*’s role in delayed wound healing, its inherent and acquired antimicrobial resistance, and the necessity for a multimodal therapeutic approach. This is due to the fact that diabetic foot infections (DFIs) are linked to diabetic foot ulcers (DFUs), which are the main cause of sepsis, and that DFIs continue to be a significant contributing factor to nontraumatic lower limb amputations, carrying a high risk of morbidity and mortality (Al-Joufi et al., 2020). The increased worry over multidrug-resistant organisms and *P. aeruginosa*’s placement in the WHO’s "critical priority pathogens" list further boosted research efforts.

##### Recent Surge (2021–2024): Resistance, Biofilms, and Targeted Interventions

Between 2021 and 2024, research activity escalated, resulting in 39 publications and peak citation rates recorded in 2024. The current period is marked by a proliferation of research on multidrug resistance (MDR) and extensively drug-resistant (XDR) strains, a focus on biofilm formation and its influence on treatment failure, molecular investigations of resistance genes and virulence factors (e.g., metallo-β-lactamases), and a vigorous pursuit of innovative therapies such as phage therapy and antimicrobial peptides. This is because globally, multidrug-resistant organisms (MDROs) such as methicillin-resistant *S. aureus* (MRSA), extended-spectrum beta-lactamase (ESBL) producers, and carbapenem-resistant Enterobacteriaceae (CRE) have drastically expanded in the last two decades (Sannathimmappa *et al.,* 2021). The increasing association between multidrug-resistant (MDR) bacteria and diabetic foot ulcers has confounded treatment approaches and heightened the risk of amputation (Hamid *et al.,* 2020). The problems of multidrug resistance (MDR), like *P. aeruginosa,* for example, have generated a serious health issue among patients with DFI (Al-Khudhairy and Al-Shammari, 2020), with their enzymes, such as metallo-β-lactamases, having the ability to hydrolyze and confer resistance to almost all β-lactam antibiotics; the lack of clinically effective metallo-β-lactamase inhibitors; the speed at which novel variants are discovered; the transferability of their encoding genes; and their ubiquity due to their isolation from nosocomial and environmental sources make them particularly interesting and concerning (Mojica *et al*., 2022). This has created the need for medications targeting this organism to increase, as most of the research output in this stage is geared towards characterizing the resistant profiles of this organism and finding a better treatment option.

#### 3.3.2 Geographic distribution of research

The distribution of research output on *Pseudomonas aeruginosa* in diabetic foot infections (DFIs) reveals that India (262 publications), the USA (238), and China (232) are the foremost contributors internationally, but Egypt (60) is the sole representative from Africa, a phenomenon attributable to several associated causes as shown in Figure 3A. Countries such as India, China, and the USA exhibit a significant prevalence of diabetes, which is associated with a heightened occurrence of diabetic foot infections and a concomitant increase in pertinent clinical and microbiological research (Saeedi et al., 2020, Aminabee *et al.,* 2024). These nations also gain from strong research infrastructure, significant financing, and access to modern diagnostic and molecular technologies, all of which enhance publication rates (OED, 2021) as empirical data demonstrates a robust association between infrastructure, financing assistance, and enhanced research productivity (Saqib and Rafique, 2021). Furthermore, public health strategies in these nations increasingly emphasize research on antimicrobial resistance (AMR) and infectious illnesses, particularly *P. aeruginosa*, owing to its clinical relevance and resistance characteristics (WHO, 2021).

**Figure 3A:**
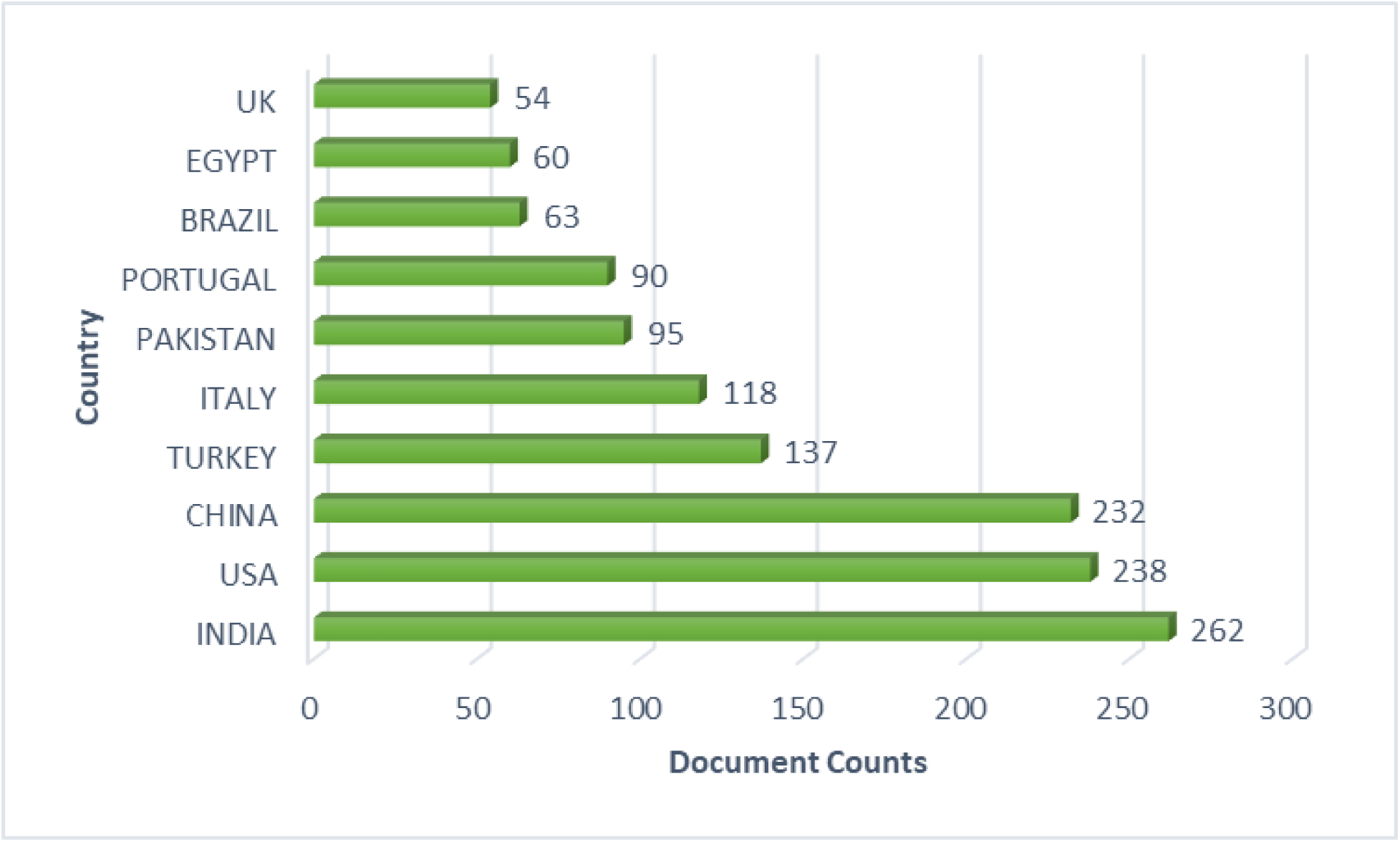
Number of publications by the top 10 countries on *Pseudomonas aeruginosa* in diabetic foot infections

**Figure 3B1:**
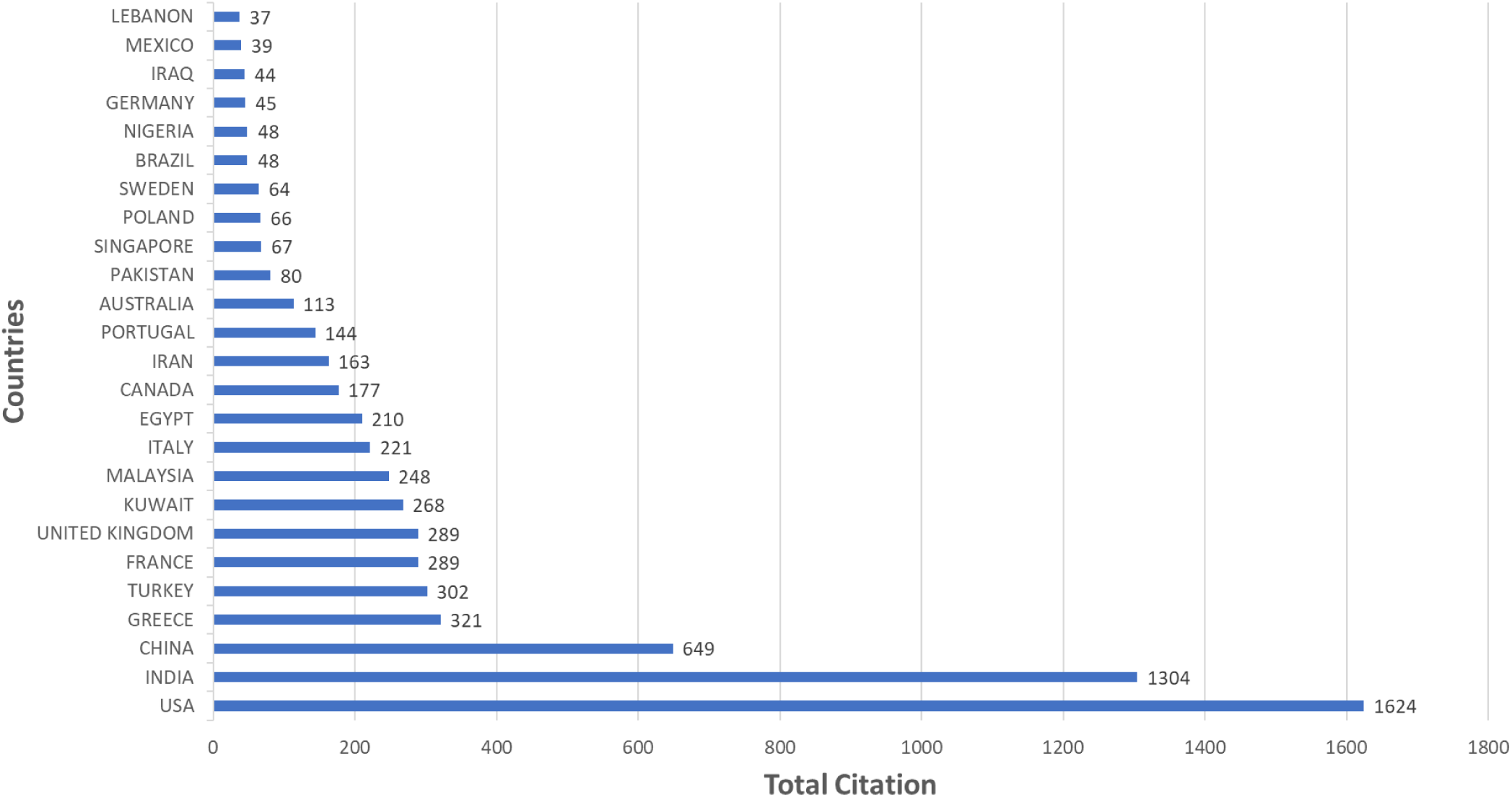
Number of Citations by Top 25 Countries on *Pseudomonas aeruginosa* in diabetic foot infections

**Figure 3B2:**
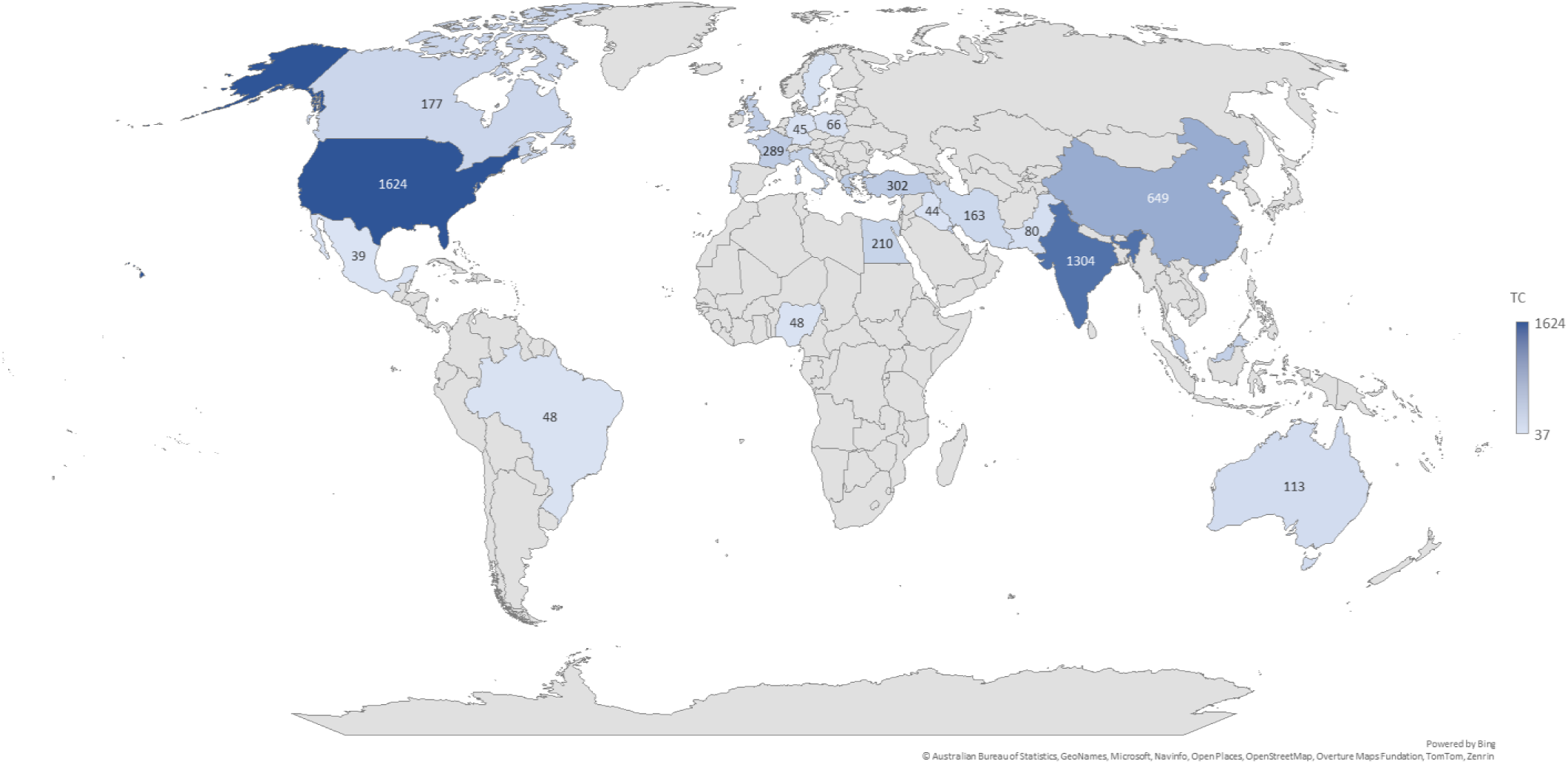
Number of Citations by Top 25 Countries on *Pseudomonas aeruginosa* in diabetic foot infections

#### 3.3.3 Most-cited documents

The most globally cited paper in this field is shown in figure 3B with the paper from Serra R. in 2015 with 486 citations, which is followed by Citron D. M. in 2007 with 369 citations, while the paper from Wolcott R. D. in 2016 had 289 citations as the third most cited paper. These studies probably filled in important gaps in our knowledge at the time they were published, especially when it came to antimicrobial resistance, polymicrobial interactions, and biofilm-related chronic wound pathology, which are all important for understanding and treating DFIs. For example, Serra et al. (2015) gave us important information about how wounds heal and how bacterial biofilms affect that process. This information is useful in many fields, including microbiology, surgery, and wound care. In the same way, Citron et al. (2007) wrote one of the first full reports on how susceptible pathogens that are often found in diabetic foot infections are to antimicrobials. This report has become a key reference in both clinical and research settings. Wolcott et al. (2016), on the other hand, helped us learn more about how chronic wounds can have more than one type of bacteria and how important it is to use precise tests when looking at the microbiome of a wound.

### 3.4 Global collaboration network trends in research on *Pseudomonas aeruginosa* in diabetic foot infections

A vibrant international collaboration network highlights the noteworthy contributions made by many nations to the scientific landscape of *P. aeruginosa* in diabetic foot infections. Since the international collaboration ratio quantifies the percentage of a nation’s research output generated in cooperation with foreign partners, this section explores nation-specific contributions to the field and looks at the complex web of international collaborations that support P. aeruginosa in diabetic foot infection research. This further demonstrates the degree of active participation in international research initiatives. Based on intra-country, or Single Country Publication (SCP), and inter-country, or Multiple Nations Publication (MCP) collaborations, Figure 4 lists the top 20 authors from each of the nations.

**Figure 4:**
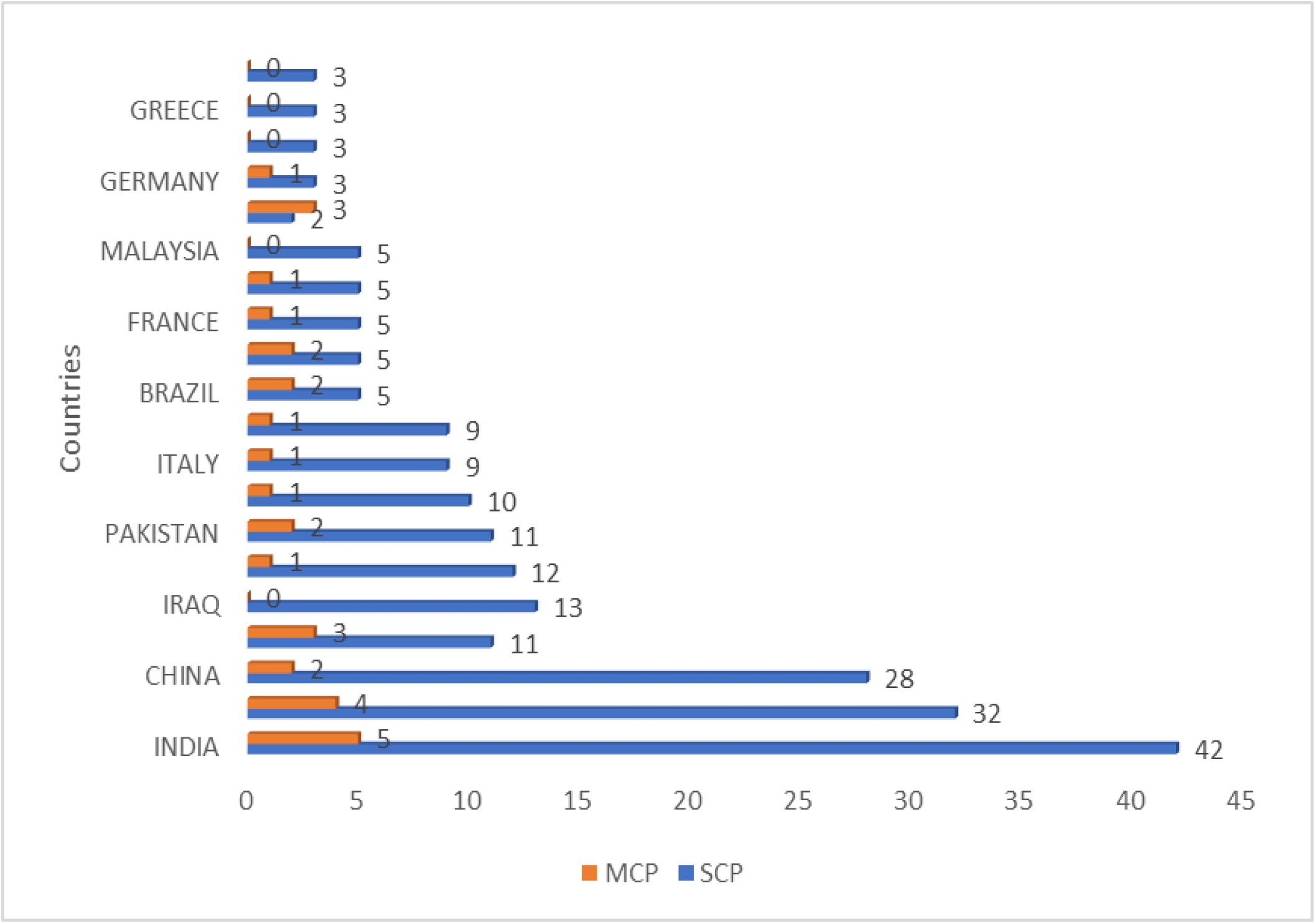
Corresponding author’s countries SCP and MCP represent intra-country and inter-country collaborations or country publications and multiple-country publications, respectively.

It was observed that intra-country publication count (SCP) was predominant in all the countries, with India having the highest SCP (SCP, 42; MCB, 5), followed by the USA (SCP, 32; MCP, 4), while China was ranked third (SCP, 28; MCP, 2). There are more single-country publications (SCP) than multi-country publications (MCP) on *Pseudomonas aeruginosa* in diabetic foot infections (DFIs) in countries like India, the USA, and China. This is a trend that is happening more and more in global collaboration networks. When SCP is dominant, it usually means that most of the research is being done within national borders, often because of funding from within the country, research priorities that are specific to that country, and strong internal institutional networks. India has a lot of SCPs, which means that its national research capacity is growing quickly. This is because the government and institutions are putting more money into health-related research, especially on diabetes and infectious diseases (Aminabee et al., 2024).

International collaborations (MCPs) usually make scientific work more visible and more likely to be cited (Gazni et al., 2012), but there may be fewer of them because of problems with logistics, administration, or funding that make it hard to work with people from other countries, especially in low- and middle-income countries. Language, political alignment, and regional health problems also play a big role in how people work together. For example, China and India have a lot of diseases and don’t need to rely on international partnerships as much because they have their own research infrastructure. This could lead to more output that is focused on their own needs (Sweileh et al., 2018). However, even though they have fewer MCPs, countries that are leaders in SCP often play important roles in regional research networks. These countries could work together more to make global research more integrated and impactful.

Furthermore, each country’s collaboration network in global research on Pseudomonas aeruginosa in diabetic foot infections (DFIs) is shown in Figure 5. This revealed 5 clusters (cluster 1-red, cluster 2-green, cluster 3-blue, cluster 4-yellow, and cluster 5-purple) with 18 links and 28 total link strengths. In Cluster 1, China had the highest number of documents (32), with 1 link and a total link strength of 1. This is followed by Pakistan with 16 documents, 4 links, and 5 total link strength, while Canada had the least number of documents (7) with 2 links and 5 total link strength. Only two countries are in cluster 2: India, which had the highest number of documents with 1 link and 2 link strength, and Saudi Arabia with 9 documents, 5 links, and 8 total link strength. Cluster 3 is made up of the United Kingdom and Turkey, which had 17 documents, 2 links, and 5 total link strengths, and Australia, which had 5 documents, 3 links, and 3 total link strengths. In Cluster 4, the USA had the highest number of documents (54) with 9 links and 15 total link strength. Brazil had 8 documents, 1 link, and 2 total link strengths, while France had 8 documents, 1 link, and 1 total link strength. Cluster 5 is made up of only Iran, with 8 documents, 2 links, and 2 total link strengths, and Iraq with 8 documents, 1 link, and 1 total link strength.

**Figure 5:**
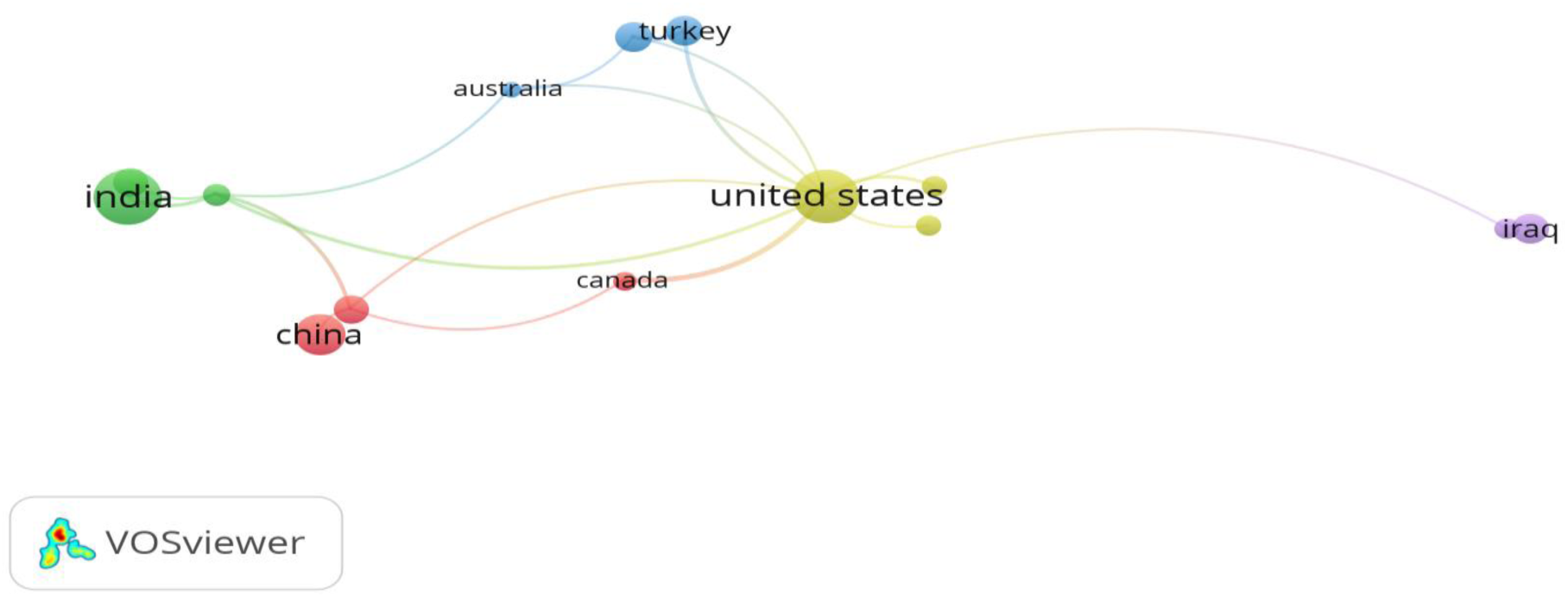
Country’s collaboration network in global research on *Pseudomonas aeruginosa* in Diabetic Foot Infection

The fact that there are five distinct clusters and relatively low link counts and total link strengths suggests that international cooperation in this area is still fragmented and focused on certain regions. The USA and China have the most publications, but they don’t work together very much. For example, China has a lot of documents but not many links, which means that most of its research is done within its own borders. This is in line with what other bibliometric studies have found: that emerging economies like China often put their own research agendas and self-reliance ahead of international co-authorship, even though they publish a lot (Zhu and Liu, 2020). On the other hand, countries like Saudi Arabia and Canada have fewer publications but stronger links, which shows that they are more engaged in working together. The same pattern can be seen with the UK and Turkey, which have moderate output but a lot of interconnectivity. These results are in line with the idea that countries with fewer resources often work together with other countries to get access to new methods, funding, and training opportunities (Gazni et al., 2012). The USA is also the leader in both the number of publications and the centrality of its network (as shown by the highest number of links and total link strength in Cluster 4). This is because the USA has always been a global research hub with many partnerships (Bornmann et al., 2015). Finding isolated clusters, like Cluster 5 (Iran and Iraq), makes it even more important to encourage more cross-regional collaborations, especially when it comes to global health priorities like antimicrobial resistance and complications from diabetes (Sweileh et al., 2018).

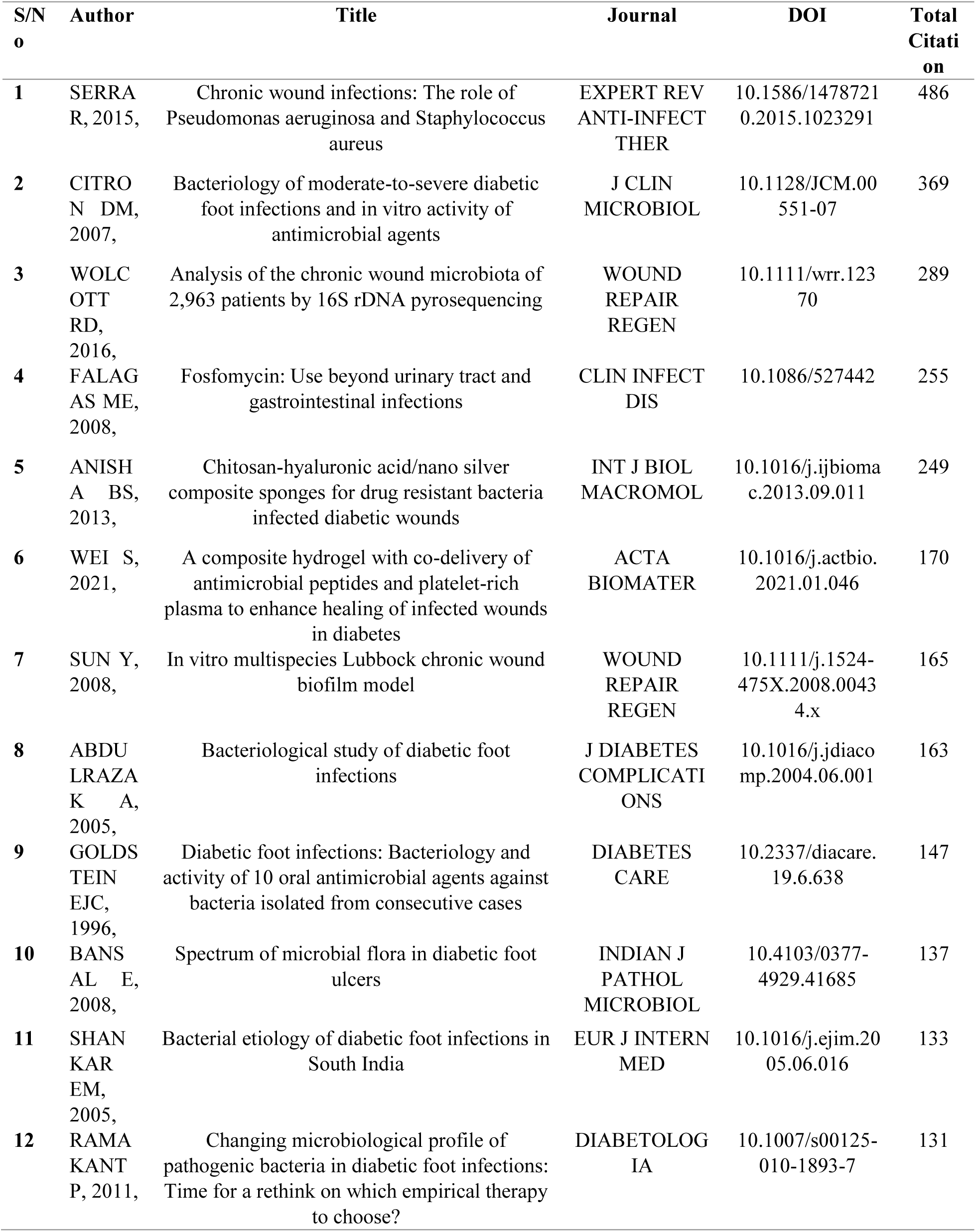
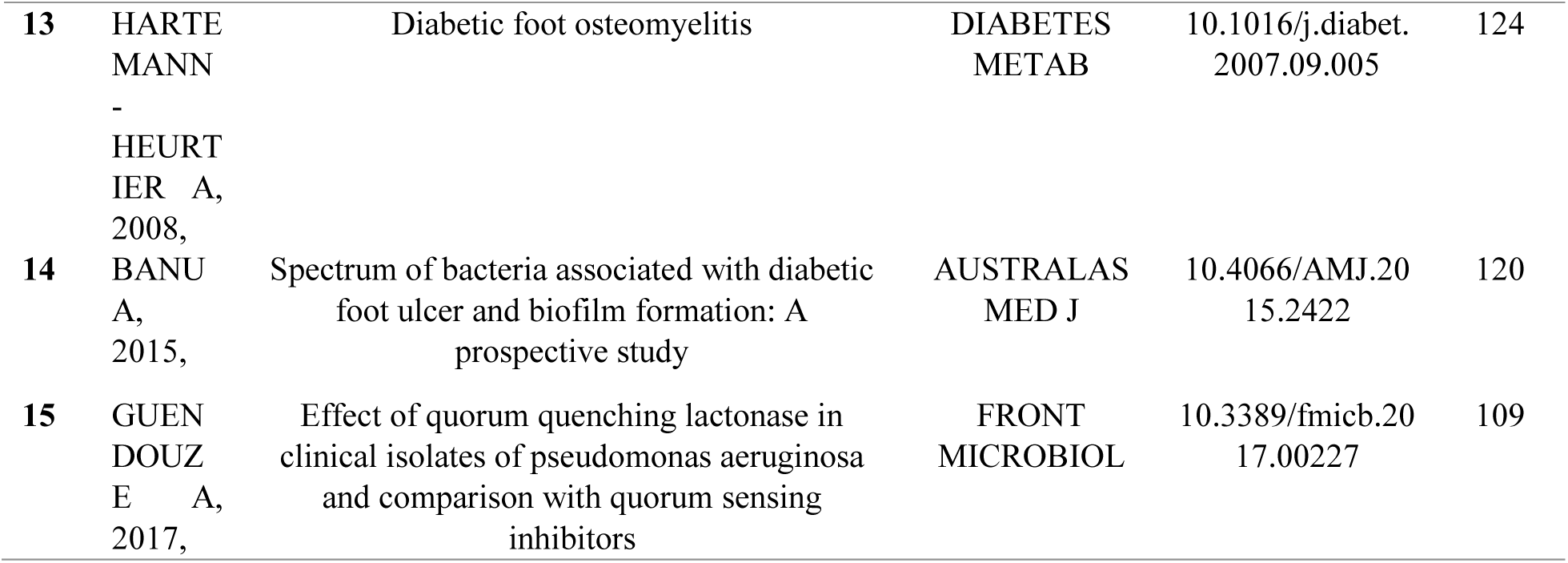

### 3.5 Descriptive statistics of relevant affiliations, academic journals, and most cited authors in research on *Pseudomonas aeruginosa* in Diabetic Foot Infection

This investigation in this research area underscores the pivotal role of prominent institutions, journals, and authors in facilitating partnerships, dissemination, and the exchange of information within the field. Moreover, these publications function as libraries of innovative research and as centers linking many lines of investigation, facilitating progress in this field. These prominent sources in this study field function as primary venues for the dissemination of unique, new discoveries and theoretical progress (Silue and Fawole, 2024).

Institutional affiliation of the 10 most relevant affiliations or institutions with the most contribution in global research on *Pseudomonas aeruginosa* in Diabetic Foot Infection are highlighted in Figure 6, with the University of Lisbon being the highest contributor of scientific articles in this field, with 30 articles. This is closely followed by Parke-Davis Pharmaceutical Research with 29 articles, while Azienda Ospedaliera Universitaria Pisana contributed 23 articles.

**Figure 6:**
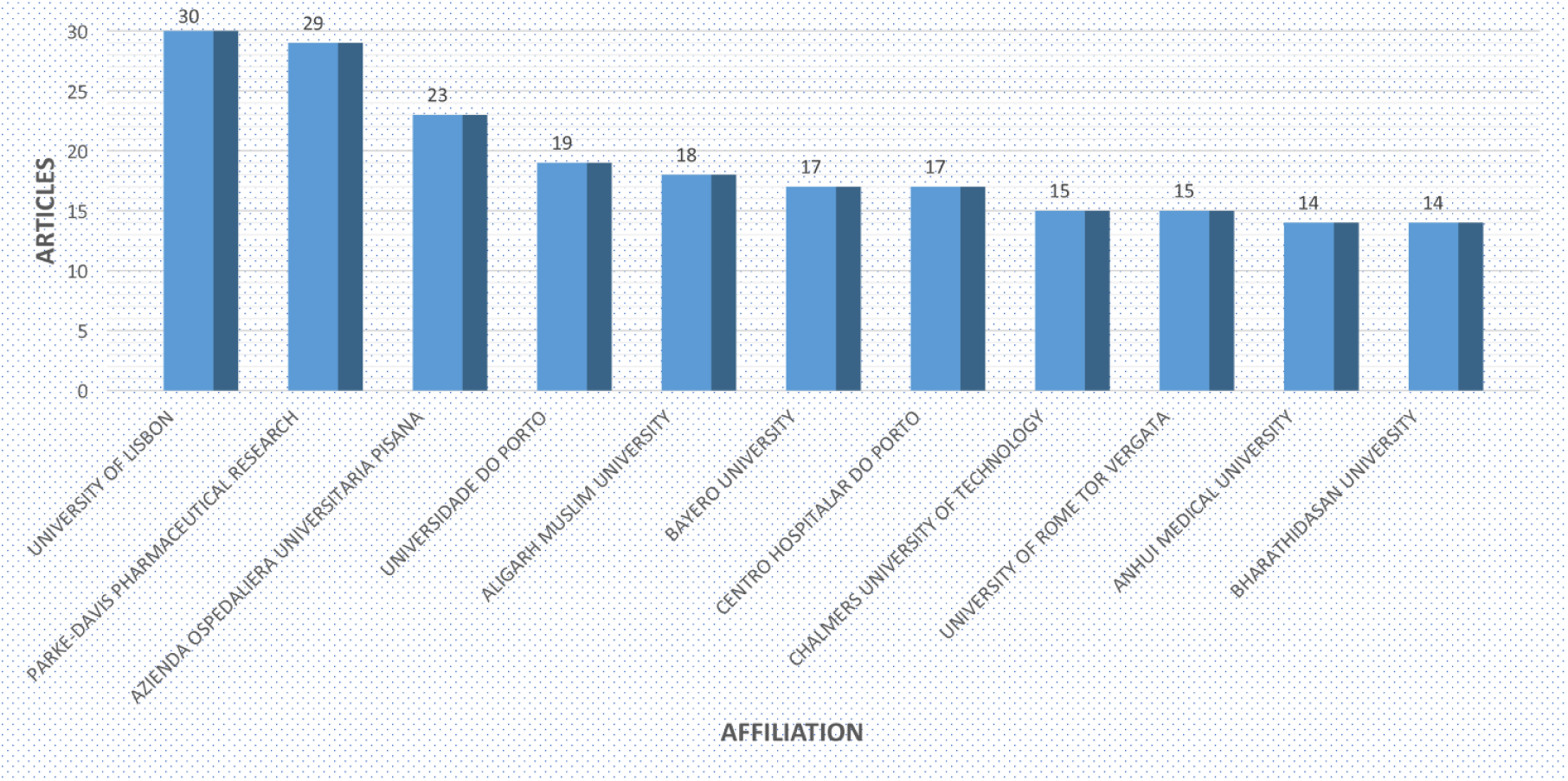
Most Relevant Affiliations in Research on *Pseudomonas aeruginosa* in diabetic foot infections

This shows that the University of Lisbon has a robust biomedical research infrastructure and is actively involved in antibiotic resistance and translational research. Recent studies on antimicrobial peptides (AMPs) as possible treatments for *P. aeruginosa* biofilms in diabetic foot infections, like Serrano et al. (2023) and Domingues et al. (2019), show how strong the university’s methods are and how well it combines high-end bioengineering platforms with clinically relevant translational studies. Parke-Davis Pharmaceutical Research, which is now part of Pfizer and has a long history, comes in second place. Its contribution shows that the pharmaceutical sector is interested in developing antibiotics and understanding how bacteria become resistant to them in chronic illnesses like DFIs caused by *P. aeruginosa*. The large production from a pharmaceutical institution underscores the therapeutic and commercial implications of drug-resistant infections in diabetic patients. Azienda Ospedaliera Universitaria Pisana, an Italian academic hospital, is famous for its multidisciplinary partnerships in infectious diseases, diabetes control, and surgical procedures. Its solid clinical background facilitates observational and interventional investigations, underlining the relevance of academics and industry in expanding knowledge and solutions.

The result in Figure 7 illustrates the top 10 relevant and productive sources publishing articles on global research on *Pseudomonas aeruginosa* in diabetic foot infection. According to sources, the International Journal of Lower Extremity Wounds had the highest volume of research output on Pseudomonas aeruginosa in diabetic foot infection (19). This is followed by Antibiotics, which had 10 articles, while Antimicrobial Agents and Chemotherapy and Journal Of Pure and Applied Microbiology had 7 articles. The International Journal of Lower Extremity Wounds is a popular source for studies on *Pseudomonas aeruginosa* in diabetic foot infections (DFIs), focusing on chronic wounds, diabetes complications, and limb preservation. Its targeted readership and theme congruence make it a desirable forum for infection management and microbiological issues in lower extremity wounds. The presence of publications like Antibiotics and Antimicrobial Agents and Chemotherapy highlights the clinical relevance of antimicrobial resistance in *P. aeruginosa* and the urgent need for improved treatment methods. This pattern of articles implies a multidisciplinary interest across wound care, microbiology, and pharmacology.

**Figure 7:**
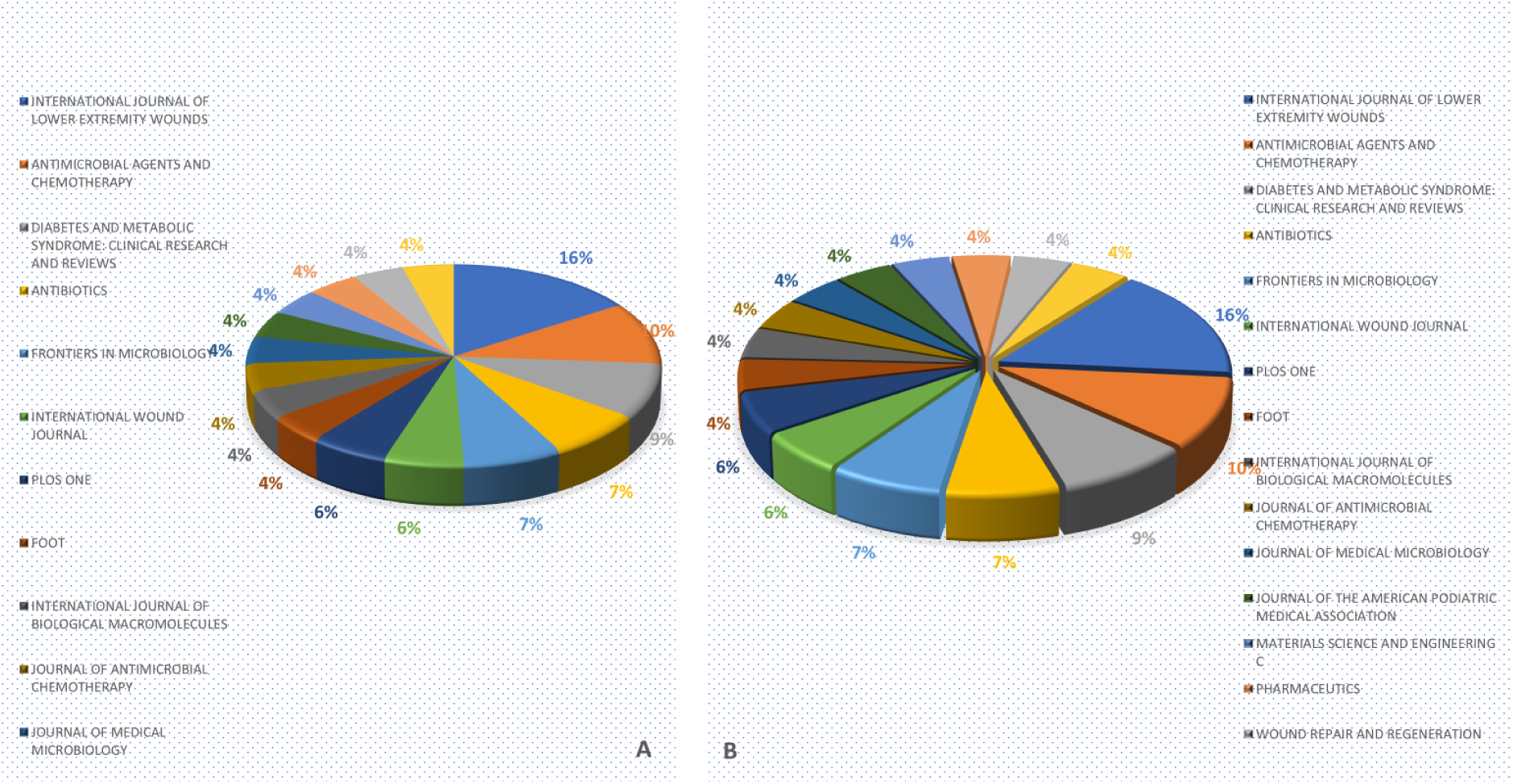
Top Relevant Sources in Global Research on *Pseudomonas aeruginosa* in diabetic foot infections. (A) Based on Impact Factor (h-index); (B) Based on Publications Counts;

The scholarly prominence and impact of journals that publish studies on *Pseudomonas aeruginosa* in diabetic foot infections (DFIs) are indicated by their H-index values. With the highest H-index of 11, the International Journal of Lower Extremity Wounds is a key publication for the dissemination of specialized information on lower limb complications and wound care for diabetic patients. Diabetes and Metabolic Syndrome: Clinical Research and Reviews (H-index 6) and Antimicrobial Agents and Chemotherapy (H-index 7) come next, both of which make substantial contributions to the general knowledge of infectious complications and metabolic control in diabetes. A targeted but expanding body of research connecting *P. aeruginosa* to DFIs within these journals is suggested by the comparatively lower H-indices in this context when compared to their global values (Resurchify, 2024; SCImago Journal Rank, 2024).

### Most productive authors

Figure 8 illustrates the trend of the top 10 authors’ productivity on global research on Pseudomonas aeruginosa in diabetic foot infections (DFIs). Lipsky, B.A., and Oleveira, M. had 7 publications, making them the most prolific authors. They are closely followed by Tavares L., who had 6 publications, while Ahmad J., Lavigne J-P., and Serrano I. had 5 publications each. Authors like Lipsky B.A. and Oleveira M. have written seven papers each on *Pseudomonas aeruginosa* in diabetic foot infections (DFIs). This shows that they have been experts in diabetic wound care and infectious disease research for a long time. Lipsky is especially well-known around the world for his work on clinical guidelines and treatment plans for DFIs, which includes research on how bacteria become resistant to drugs and how to treat them. Authors like Tavares L., Ahmad J., Lavigne J-P., and Serrano I. have also made important contributions to our understanding of wound infections from both a microbiological and clinical point of view. They have done this by studying things like biofilm formation, antimicrobial susceptibility, and translational therapeutics. The high level of research productivity of these people shows how important it is to use an interdisciplinary approach that includes microbiology, clinical medicine, and pharmacology to deal with *P. aeruginosa*-related problems in diabetic patients. Their repeated appearances in the literature also suggest that there are active institutional support and international collaboration networks that make it possible for people to keep publishing.

**Figure 8:**
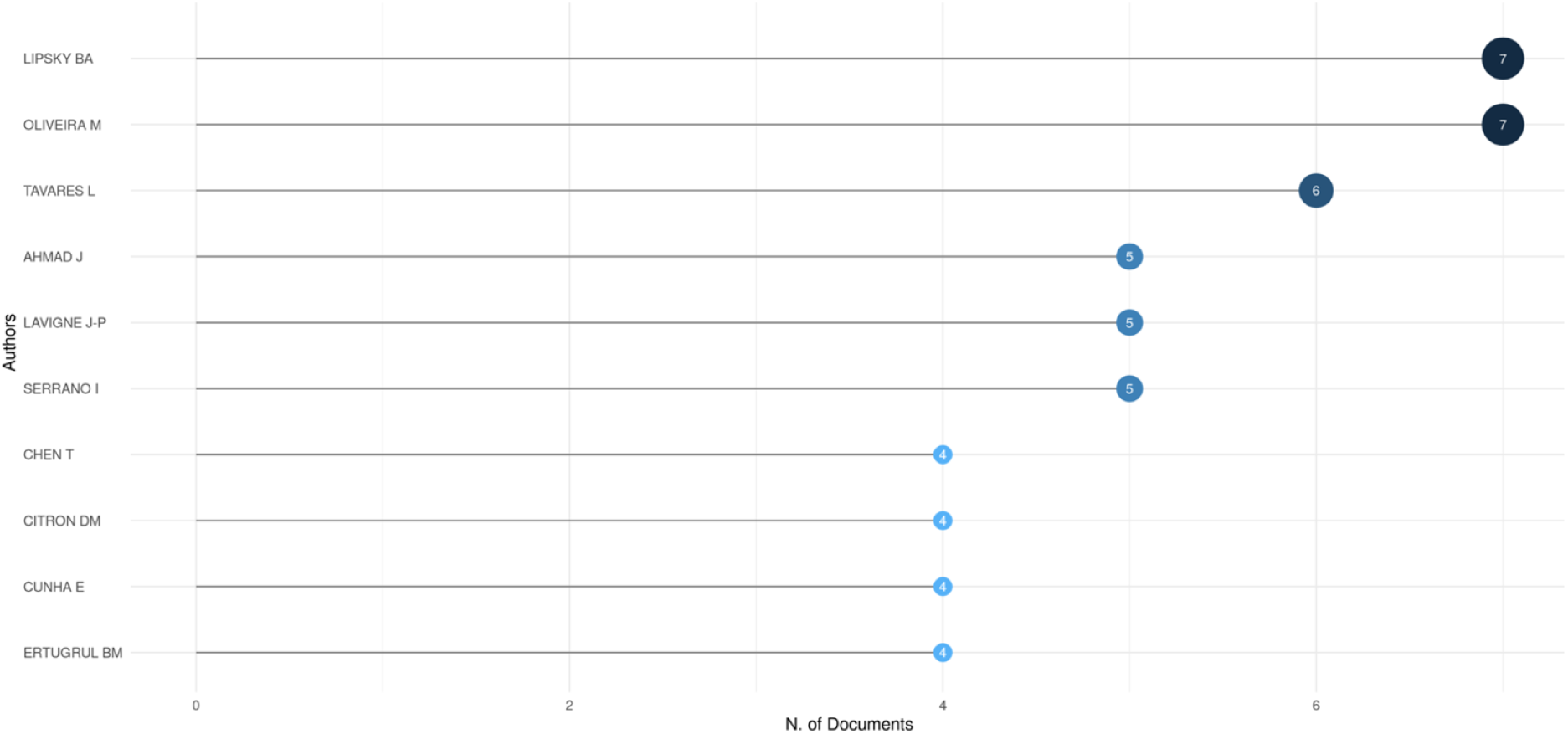
The Most Relevant Authors in Global Research on *Pseudomonas aeruginosa* in diabetic foot infections.

**Figure 9:**
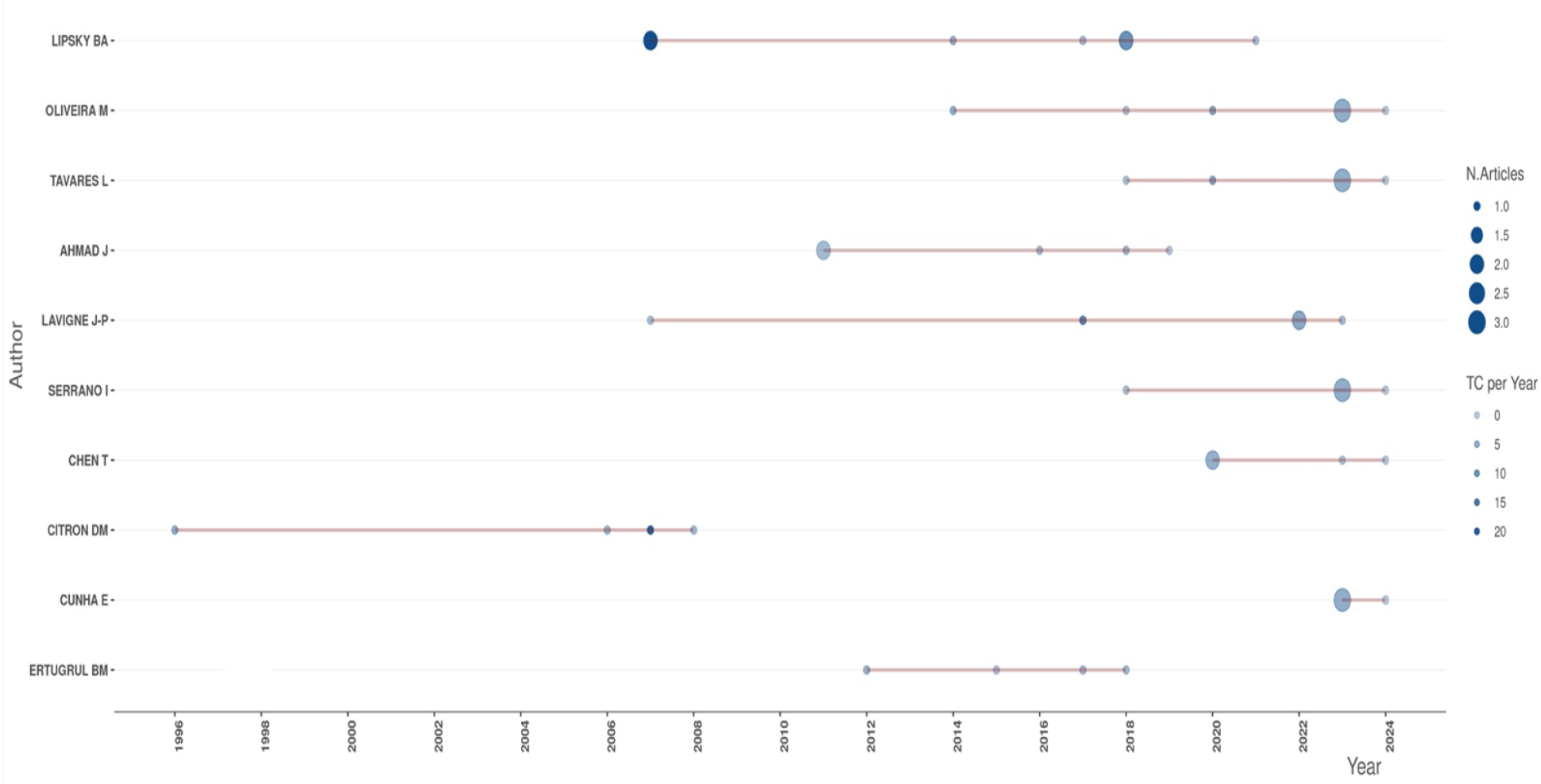
Top 10 Authors’ Productivity Global Research on *Pseudomonas aeruginosa* in diabetic foot infections Over Time from 1996 to 2024.

Authors production over time revealed that Lipsky, B.A., a significant contributor in the field, made his highest contribution to the field of study in 2007 and 2018 with 2 articles, while in 2014, 2017, and 2021, he produced one article in each of these years. Oliveira’s peak contribution was 2023, with 3 publications, while 2014, 2018, 2020, and 2024 produced one article each. Tavares L.’s peak contribution was in 2023 with 3 publications. This has come to prove the contributions of these authors in shaping this field of study and their significant influence in this field of study.

**Table 4:**
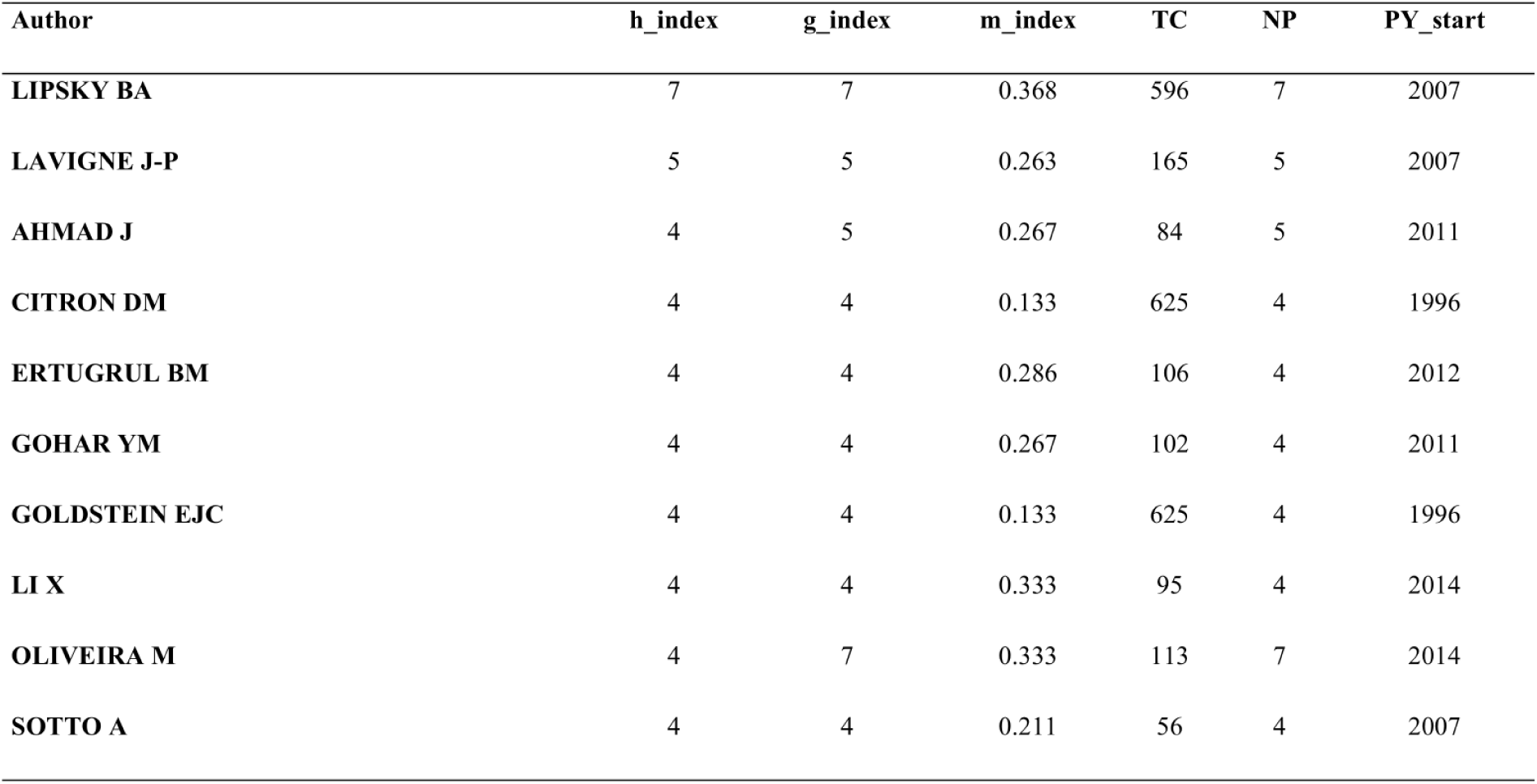
Top 10 Authors Global Research on *Pseudomonas aeruginosa* in diabetic foot infections.

| Author | h_index | g_index | m_index | TC | NP | PY_start |
| --- | --- | --- | --- | --- | --- | --- |
| LIPSKY BA | 7 | 7 | 0.368 | 596 | 7 | 2007 |
| LAVIGNE J-P | 5 | 5 | 0.263 | 165 | 5 | 2007 |
| AHMAD J | 4 | 5 | 0.267 | 84 | 5 | 2011 |
| CITRON DM | 4 | 4 | 0.133 | 625 | 4 | 1996 |
| ERTUGRUL BM | 4 | 4 | 0.286 | 106 | 4 | 2012 |
| GOHAR YM | 4 | 4 | 0.267 | 102 | 4 | 2011 |
| GOLDSTEIN EJC | 4 | 4 | 0.133 | 625 | 4 | 1996 |
| LI X | 4 | 4 | 0.333 | 95 | 4 | 2014 |
| OLIVEIRA M | 4 | 7 | 0.333 | 113 | 7 | 2014 |
| SOTTO A | 4 | 4 | 0.211 | 56 | 4 | 2007 |

### 3.6 Emerging trends and research gaps in global research on *Pseudomonas aeruginosa* in Diabetic Foot Infection

#### 3.6.1 Citation network analysis - Trending topics

Figure 10 illustrates trending topics in the research area on *P. aeruginosa* in diabetic foot infection. From the early 2000s to 2024, there was a noticeable shift in thematic focus, according to the Trend Topics analysis of international research on *Pseudomonas aeruginosa* in diabetic foot infections. Terms like heat, cellulitis, penicillin G, and bacteria, anaerobic, were highlighted in early research. More clinically and microbiologically specific themes developed over time, with terms like wound infection, *Pseudomonas aeruginosa*, antibiotic resistance, microbiology, and bacterium isolation becoming more common, particularly after 2016. Notably, diabetes mellitus, diabetic foot ulcers, and epidemiology have taken center stage in recent years, reflecting a growing concern for managing chronic wounds and antibiotic resistance on a global scale. The term frequency bubbles also show that subjects like antibiotic resistance and diabetic foot ulcers were among the most researched, indicating ongoing interest in these fields and their potential for clinical application. This pattern highlights a move toward interdisciplinary research that combines public health, clinical management, and microbiology.

**Figure 10:**
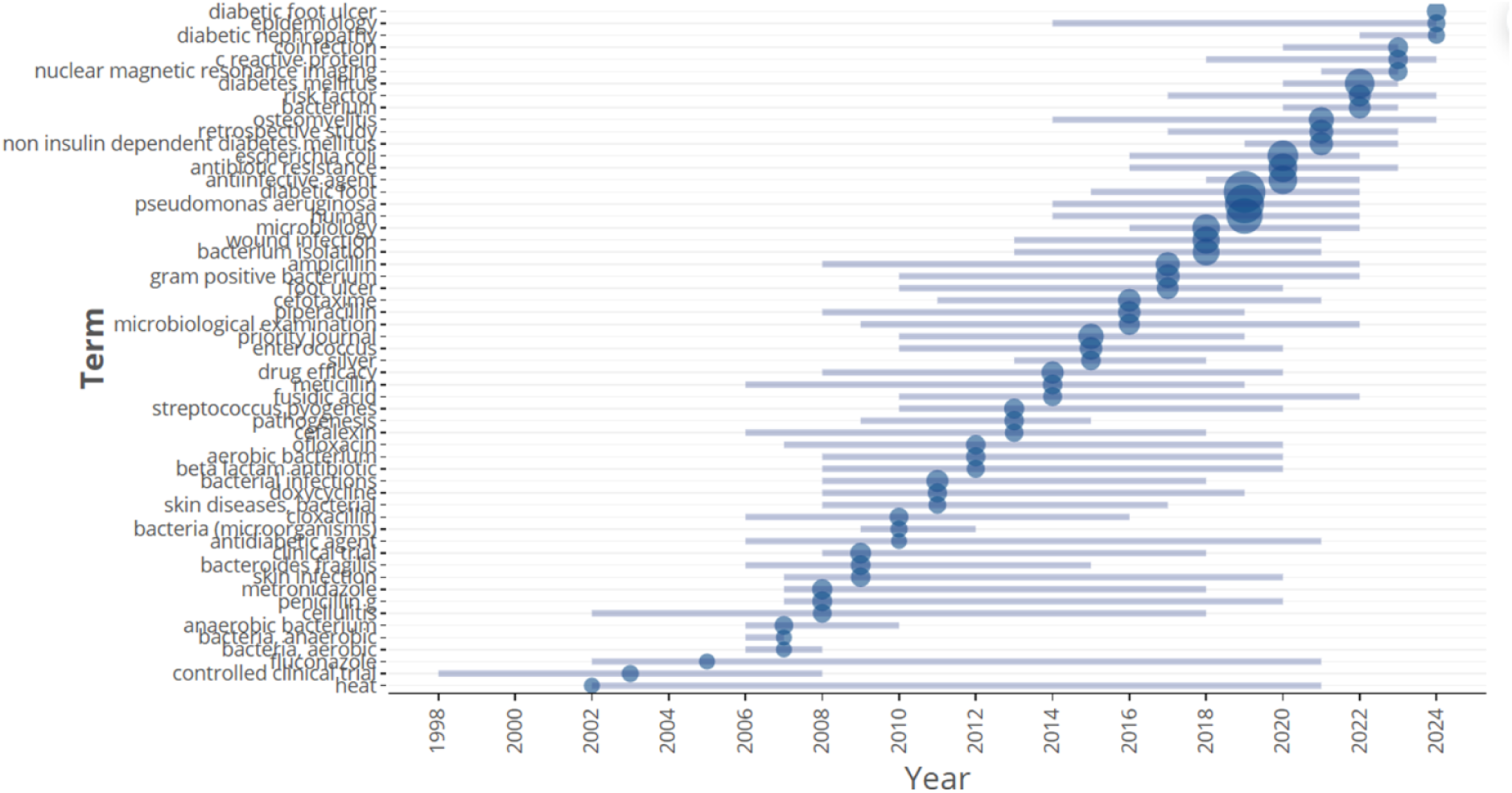
Trending Topics in Global Research on *Pseudomonas aeruginosa* in Diabetic Foot Infections.

#### 3.6.2 Thematic map

The conceptual structure of research on Pseudomonas aeruginosa in diabetic foot infections (DFIs) is visualized by the thematic map, which is depicted in Figure 11 and is based on centrality (relevance) and density (development). As the fundamental focus of the field, the fundamental themes in the bottom-right quadrant—human, *Pseudomonas aeruginosa*, and diabetic foot—are extremely pertinent but have not yet reached their full potential. Male, female, and adult motor themes are central and well-developed in the upper-right quadrant, suggesting that demographic factors are important and being studied. Terms like vancomycin, ciprofloxacin, gentamicin, and amikacin, which are represented in the niche themes quadrant (top-left), indicate specialized interest in antibiotic therapy because they are specific, highly developed topics with little connection to the wider field. Animal experiments, biofilm, in vitro research, and antibacterial activity are the emerging or waning themes in the bottom-left quadrant. These themes exhibit low development and relevance, suggesting that they are either understudied or losing interest. The maturity and areas of focus of current scientific research in this field are revealed by this thematic distribution.

**Figure 11:**
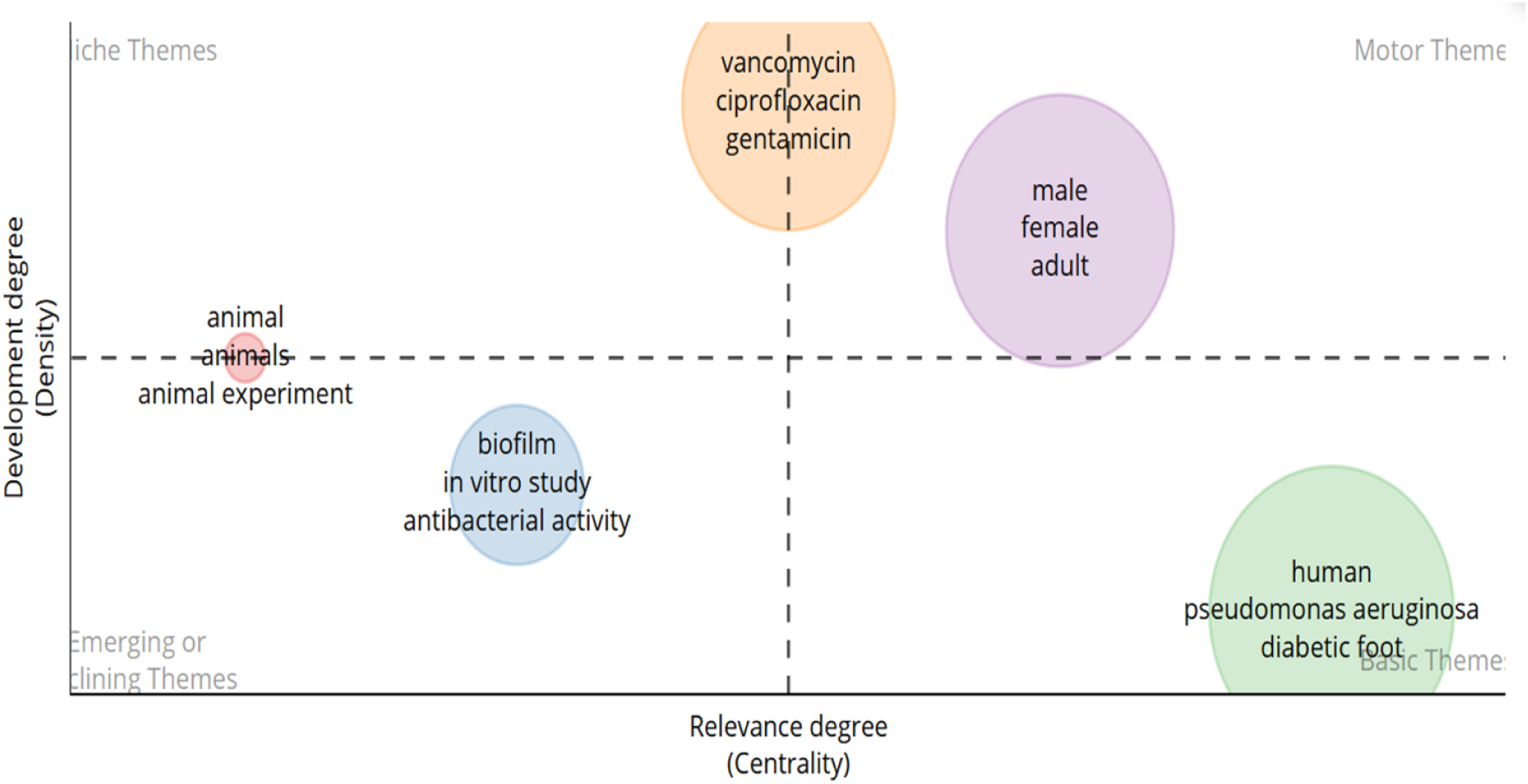
Thematic map from keywords plus in Global Research on *Pseudomonas aeruginosa* in Diabetic Foot Infections.

#### 3.6.3 Thematic evolution

Figure 12 displays the thematic evolution map that charts the research trends concerning diabetic foot complications between 1985 and 2024. Three main time periods are distinguished in the timeline: 1985–2013, 2014–2021, and 2022–2024. General themes such as diabetes, diabetic foot, infections, and *Staphylococcus aureus* were the focus of early research (1985–2013). From 2014 to 2021, focus shifted to more specialized clinical and microbiological problems like MRSA, diabetic foot ulcers, antibacterial activity, and multidrug-resistant bacteria. As interest in pathogen-specific difficulties and resistance mechanisms in the treatment of diabetic foot ulcers has grown, the focus has shifted to more complex issues in the most recent period (2022–2024), such as biofilm formation, antimicrobial resistance, and *Pseudomonas aeruginosa*. The map shows a distinct shift from general foundational research to themes that are more in-depth, pathogen-specific, and resistance-related. The thematic evolution map shows how research topics pertaining to diabetic foot complications have evolved chronologically, showing how scholarly attention has shifted over time from broad to more intricate and specialized issues. Between 1985 and 2013, the majority of research focused on general topics like diabetes, diabetic foot disorders, and infections, with a focus on common pathogens like *Staphylococcus aureus*. The basis for comprehending the clinical consequences of diabetic foot was established by this foundational stage (Boulton et al., 2005). The emphasis shifted to more complex topics between 2014 and 2021, including multidrug-resistant bacteria, diabetic foot ulcers (DFUs), and antibacterial and antimicrobial reactions (Lipsky et al., 2016). Concern over infection control and resistance trends grew during this time. *Pseudomonas aeruginosa* infections, which are notoriously difficult to treat in DFUs, biofilm formation, and antimicrobial resistance are among the advanced microbiological and resistance-related issues that have been addressed in the most recent period, 2022–2024 (Ramirez-Acuña et al., 2019). The need for targeted therapeutic approaches and growing clinical complexity are reflected in this evolution, underscoring the need to address drug resistance and enhance wound care results.

**Figure 12:**
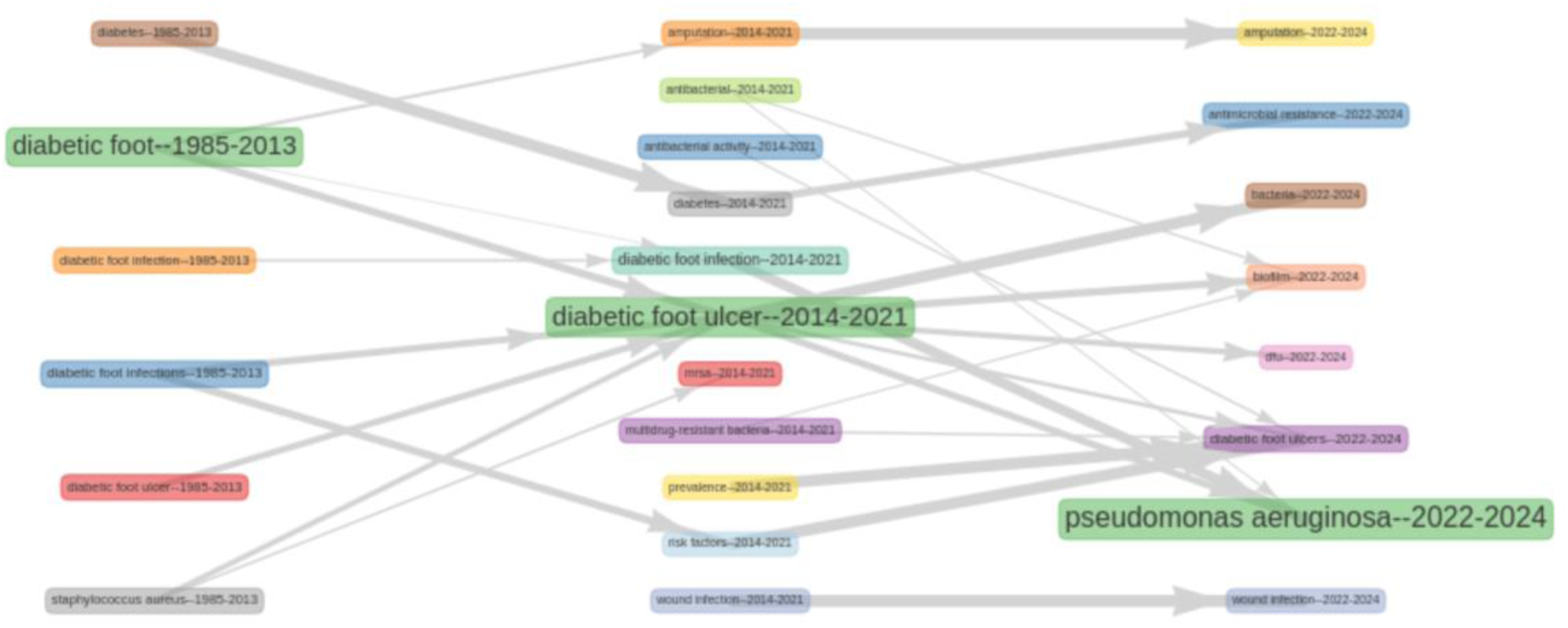
Thematic evolution of author keywords in global research on *Pseudomonas aeruginosa* in Diabetic Foot Infection.

## 4.0 Co-occurrence network analysis

### 4.1 Co-occurrence—all keywords

Figure 13 displays a visual network map of commonly co-occurring keywords used in scientific literature, representing the entire keyword co-occurrence network in worldwide research on *Pseudomonas aeruginosa* in diabetic foot infection. The main research themes and their connections within the field are highlighted by this co-occurrence analysis. Important terms that play a central role in the discourse include *Pseudomonas aeruginosa*, diabetic foot infection, biofilm, antibiotic resistance, wound healing, multidrug resistance, and chronic wounds. Different but related thematic areas, including microbiology, clinical management, antimicrobial strategies, and molecular mechanisms, are suggested by the clustering of keywords. In the study of *P. aeruginosa* and its effect on diabetic foot complications, the network visualization aids in identifying research priorities, new trends, and interdisciplinary connections. Core terms that highlight *P. aeruginosa*’s clinical relevance as a persistent and treatment-resistant pathogen in chronic wounds include biofilm, antibiotic resistance, wound healing, and multidrug resistance (Ramirez-Acuña et al., 2019). The terms’ co-occurrence highlights the pathogen’s role in delayed healing and the increasing problem of antimicrobial resistance, reflecting interdisciplinary concerns spanning microbiology, pharmacology, and wound care (Lipsky et al., 2020). The necessity of integrated therapeutic approaches and ongoing research into focused interventions for *P. aeruginosa* in DFIs is highlighted by this network analysis.

**Figure 13:**
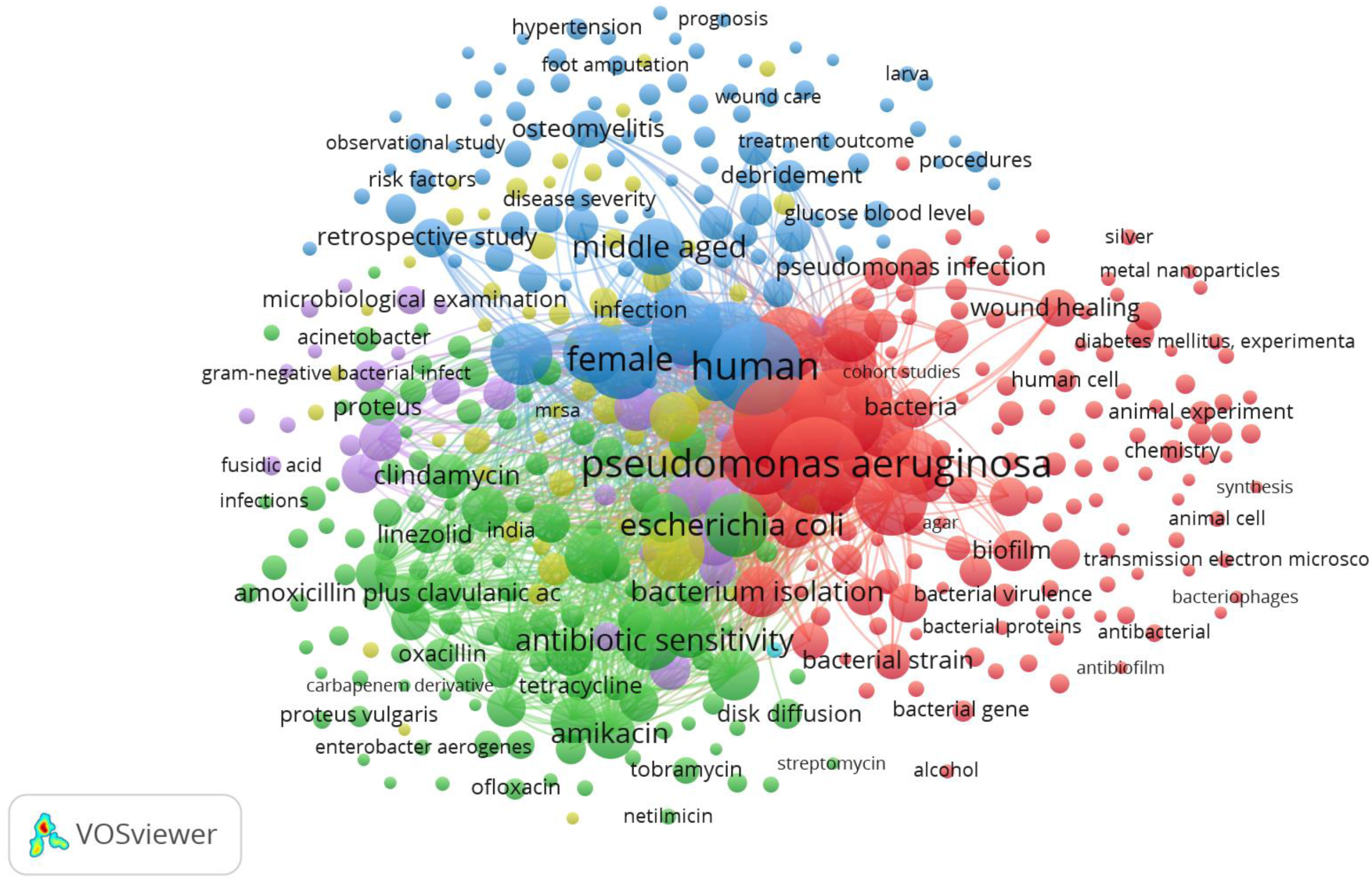
All keywords co-occurrence network in global research on *Pseudomonas aeruginosa* in Diabetic Foot Infection.

### 4.2 Co-occurrence authors keywords

Figure 14 shows the authors’ keyword co-occurrence network, which highlights the main thematic clusters in the world’s research on *Pseudomonas aeruginosa* in diabetic foot infections (DFIs). The network reveals a number of noteworthy clusters. The Yellow Cluster is dominated by *Pseudomonas aeruginosa* and is closely associated with *Staphylococcus aureus*, chronic wounds, wound infection, and antibacterial strategies, indicating microbial diversity and wound healing complications; the Blue Cluster highlights diabetic foot, including terms like "foot ulcer," "amputation," and "microbiology," representing the clinical and diagnostic dimensions of DFIs; and the Purple Cluster centers on diabetic foot infection, linking it with *P. aeruginosa*, antimicrobial resistance, and diabetes, stressing infection pathology and microbial resistance. Red Cluster links diabetes mellitus, polymicrobial infection, prevalence, and antibiotic susceptibility, indicating underlying conditions and epidemiological concerns; Green Cluster includes terms such as diabetic foot ulcers, osteomyelitis, antibiotics, and antibacterial activity, reflecting treatment approaches and complications. With *P. aeruginosa* as a primary focus because of its role in persistent and drug-resistant infections, this network illustrates how research on DFIs has expanded to include multidisciplinary perspectives, including microbiology, resistance mechanisms, clinical outcomes, and therapeutic interventions (Lipsky et al., 2020; Ramirez-Acuña et al., 2019).

**Figure 14:**
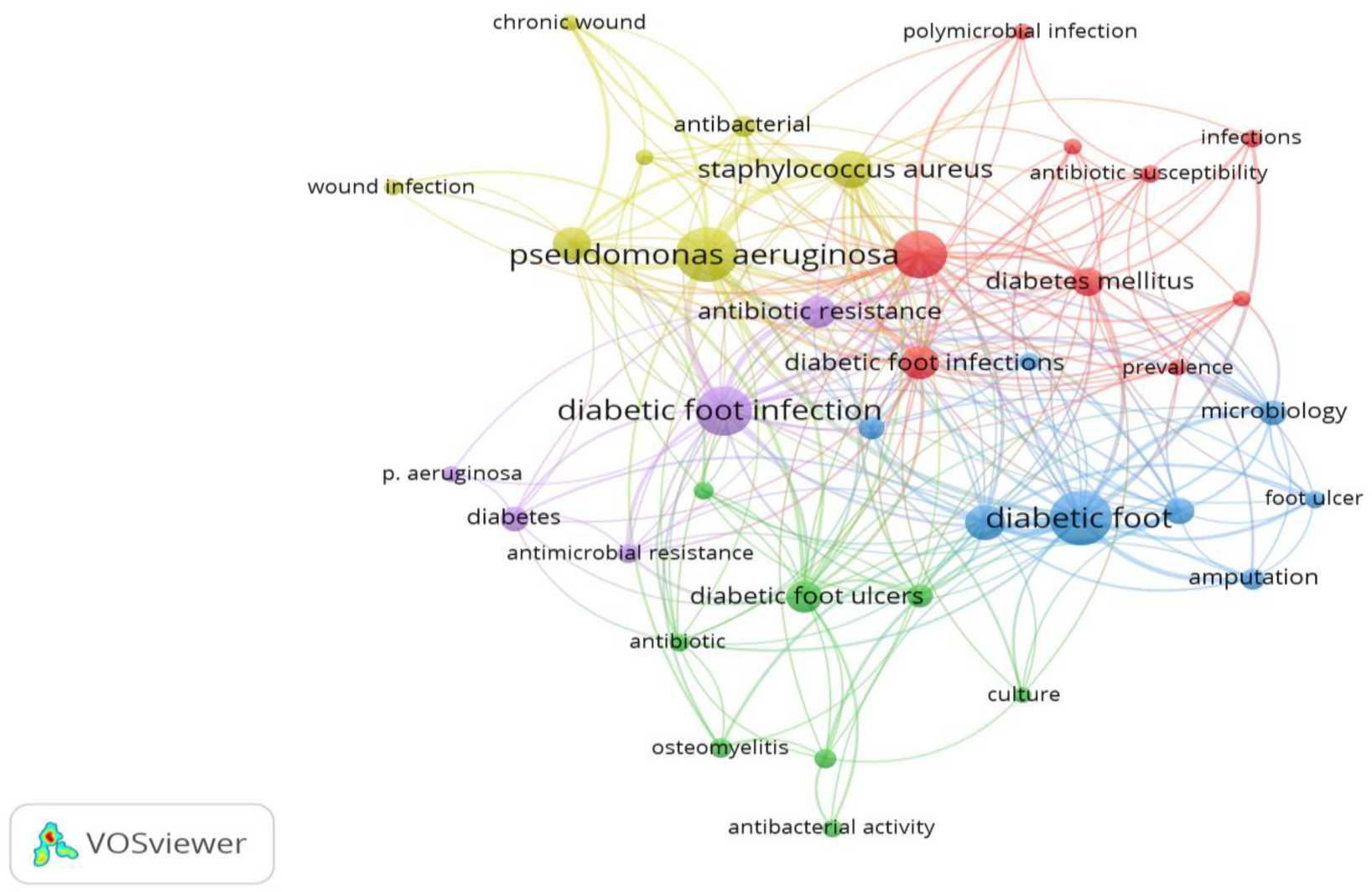
Author keywords co-occurrence network on global research *Pseudomonas aeruginosa* in Diabetic Foot Infection

### 4.3 Co-occurrence-index keywords

Five major keyword clusters that define worldwide research on Pseudomonas aeruginosa in diabetic foot infections (DFIs) are revealed by the co-occurrence index keyword in figure 15, each of which represents a different thematic focus. The clinical spectrum and diagnostic aspects of diabetic foot infections (DFIs) are represented by the Blue Cluster: Clinical Features and Management. This cluster includes terms such as "clinical trials," "middle-aged patients," "skin infections," and "outcome assessments," suggesting an ongoing focus on patient-centered studies. These keywords draw attention to the frequency, seriousness, and involvement of diabetic foot complications in conditions that pose a threat to limbs (Lipsky et al., 2020). The yellow cluster indicates interdisciplinary research linking clinical practice and molecular microbiology by bridging both clinical and pharmacological themes, such as diabetic foot infection and beta-lactamases. Green Cluster: Mechanisms of Resistance and Treatment: This cluster focuses on therapeutic interventions and challenges in treating deep-tissue infections and biofilm-associated bacteria, and it includes terms like diabetic foot ulcers, osteomyelitis, antibiotics, antibacterial activity, and antimicrobial resistance (Lipsky et al., 2020). In order to comprehend bacterial pathogenesis and evaluate treatment strategies, the red cluster concentrates on laboratory and experimental research on wound healing, biofilm formation, chronic wounds, and the use of animal models. All of these clusters demonstrate how research is becoming more interdisciplinary in order to address the complexity of DFIs and the growing threat of antibiotic resistance in *Pseudomonas aeruginosa*. This interdisciplinary approach links microbiological insight with clinical management.

**Figure 15:**
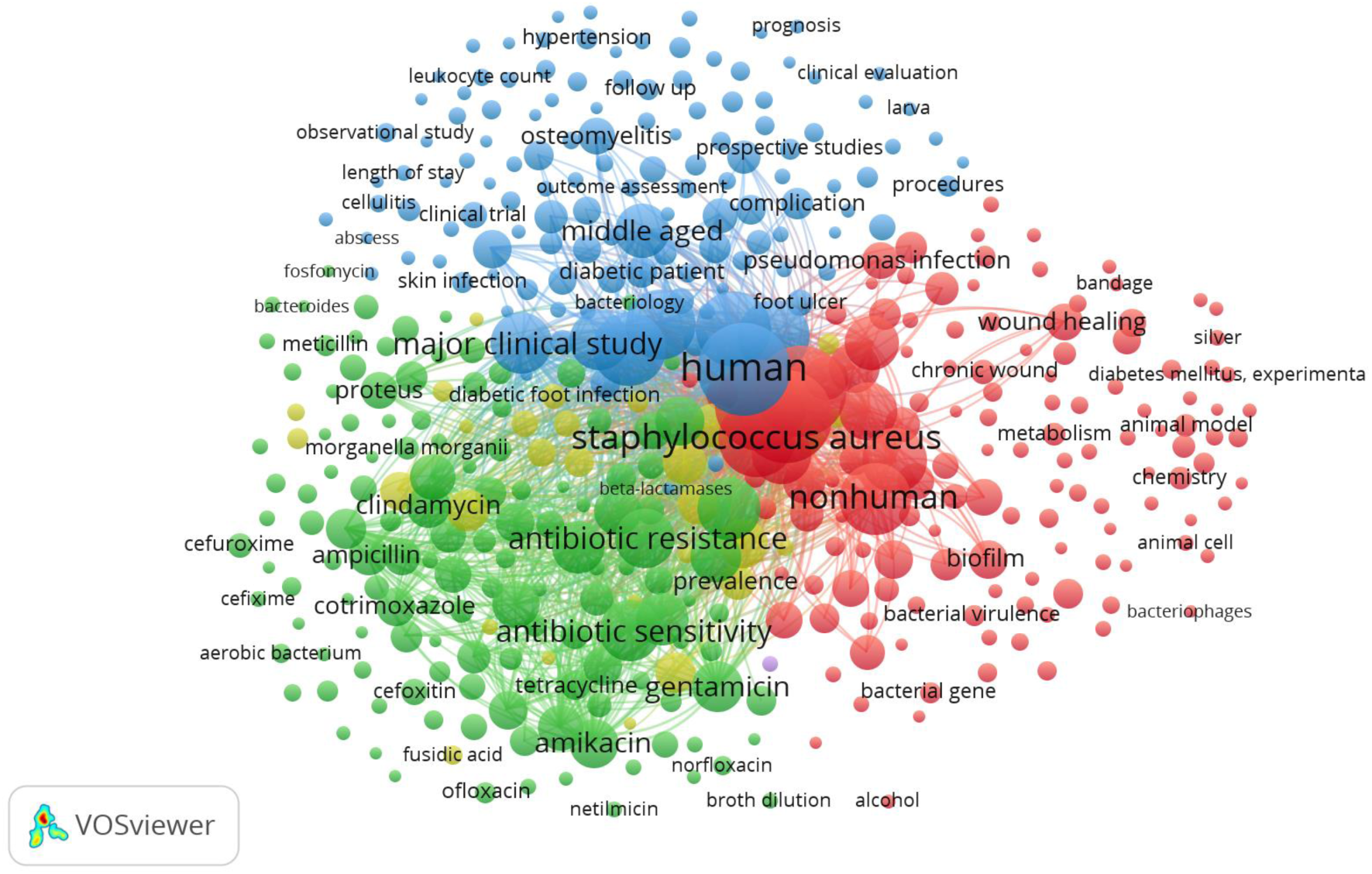
Index keywords co-occurrence overlay in global research on global research *Pseudomonas aeruginosa* in Diabetic Foot Infection

## 5.0 Network of the co-authorship countries

Analyzing co-authorship countries in bibliometric studies is crucial for understanding global collaboration patterns and the geographical distribution of scientific knowledge production. It helps identify influential countries, their capacity for international cooperation, funding, infrastructure, and scientific impact. This analysis helps assess international partnerships, detect regional research hubs, and identify potential disparities in scientific contributions. In the context of *Pseudomonas aeruginosa* and diabetic foot infections, it can guide future collaborations, support resource allocation, and promote equitable global research participation.

Based on co-authored publications on international research on *Pseudomonas aeruginosa* in Diabetic Foot Infection, Figure 16 shows the patterns of international collaboration among different nations. The United States is the most visible and well-connected node at the network’s core, indicating its leading position in international research cooperation. Different color-coded clusters that each represent groups of nations with strong internal cooperation are visible in the visualization. The United States, France, and Brazil are included in the yellow cluster, which suggests a direct but restricted research collaboration between these nations. China, Canada, and Pakistan, on the other hand, belong to the red cluster and exhibit connections to the United States as well as co-authorship relationships. India’s growing involvement in international research is demonstrated by the green cluster’s strong collaborative ties with Saudi Arabia and the United States, both in the green cluster. With further ties to the United States, the blue cluster—which includes Australia, the United Kingdom, and Turkey—reflects regional cooperation. Iraq and Iran, which are represented by the purple cluster in the dataset, seem to be comparatively isolated and only have a connection to the United States, indicating little interaction with other nations. All things considered, the node sizes and link thicknesses highlight that the US is the primary hub in this co-authorship network, with China and India emerging as significant secondary collaborators.

**Figure 16:**
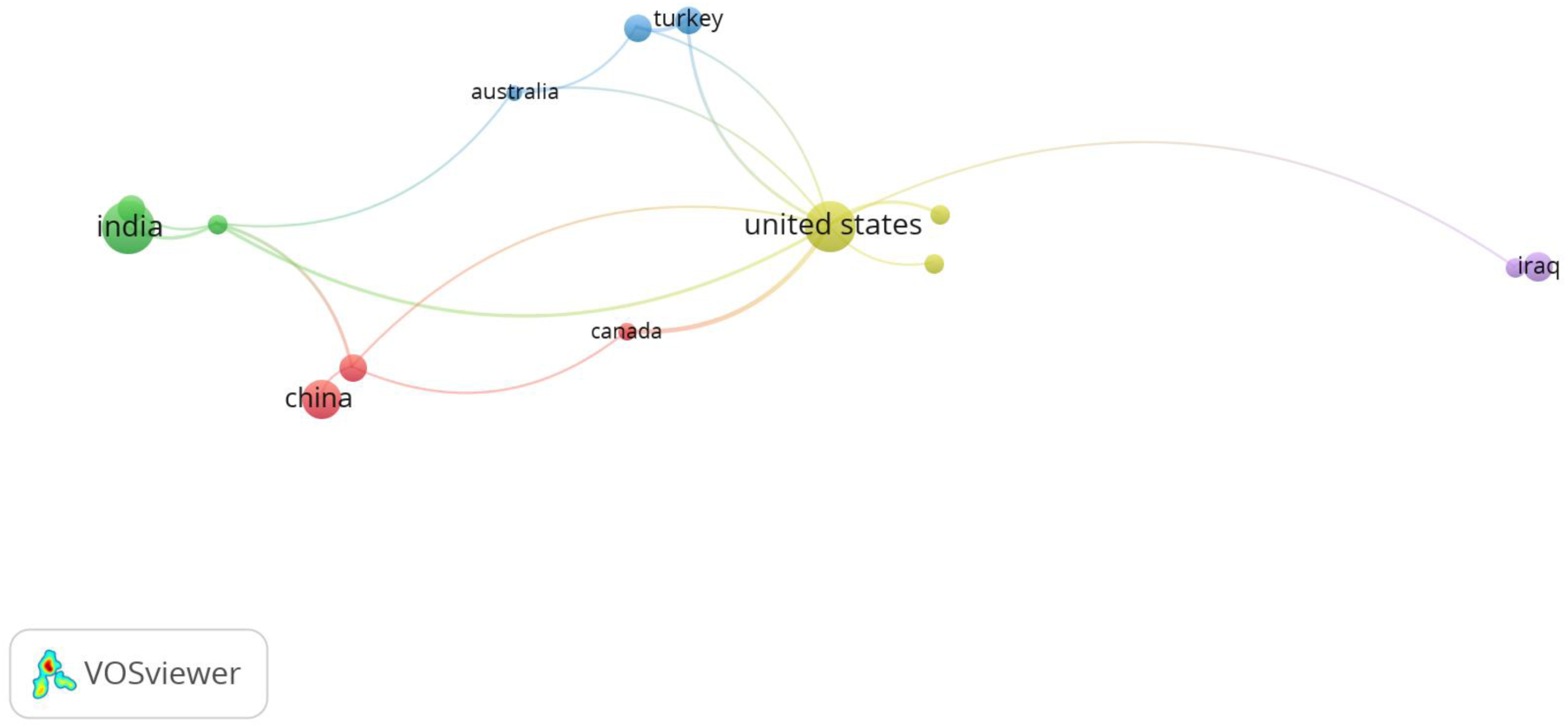
Network of the co-authorship countries in global research on *Pseudomonas aeruginosa* in Diabetic Foot Infection

## 6.0 Discussion

### 6.1 Global research on *P. aeruginosa* evolved in focus and intensity over the last two decades

The amount and thematic complexity of research on Pseudomonas aeruginosa in diabetic foot infections (DFIs) have increased dramatically over the past 20 years. After a modest start in the 1980s and intermittent attention in the early 1990s, the field saw a significant increase in scientific output starting in 2005. This was fueled by the growing clinical challenge posed by multidrug-resistant strains of P. aeruginosa and the increasing prevalence of chronic wounds related to diabetes (Garousi et al., 2023; Lee et al., 2023). Globally, research is shifting toward pathogen-specific studies, with a growing emphasis on *P. aeruginosa*’s virulence factors, biofilm-forming abilities, and designation as a WHO priority pathogen (De Oliveira et al., 2020; WHO, 2021). The most productive phase, with a peak in publications and citations, occurred between 2021 and 2024. This was in line with developments in molecular diagnostics, the appearance of resistance mechanisms like metallo-β-lactamases, and the investigation of new therapeutic approaches like phage therapy and quorum sensing inhibitors (Mojica et al., 2022; Serrano et al., 2023). A shift from broad discussions of infection and wound care to a multidisciplinary focus combining microbiology, pharmacology, and public health is revealed by thematic evolution analyses indicating ongoing clinical and scholarly interest in this resilient pathogen. Keywords such as "biofilm," "antibiotic resistance," and "chronic wounds" have taken center stage (Ramirez-Acuña et al., 2019; Lipsky et al., 2020).

### 6.2 Emerging subdomains as hotbeds of innovation

Numerous subdomains have recently surfaced as hubs of innovation among the many thematic areas investigated in Pseudomonas aeruginosa research pertaining to diabetic foot infections (DFIs).

The organism’s capacity to create intricate, robust biofilm structures that protect it from host immune responses and antimicrobial agents, thereby making treatment considerably more difficult, is the main reason why biofilm research has drawn more attention recently (Garousi et al., 2023; Rhoads et al., 2013). The comprehension of biofilm-regulated virulence factors, including pyocyanin, proteases, and rhamnolipids, which are frequently regulated by complex quorum-sensing systems, has become more important as a result (Chichón et al., 2023). Since biofilms are now known to play a major role in chronicity and delayed wound healing in DFIs, new therapeutic approaches are being developed to target them.

Concurrently, the emergence of antimicrobial resistance (AMR), particularly in multidrug-resistant (MDR) and extensively drug-resistant (XDR) strains, has sparked concern worldwide and increased research into acquired and intrinsic resistance mechanisms, including β-lactamases, efflux pumps, and metallo-β-lactamase enzymes (De Oliveira et al., 2020; Mojica et al., 2022). Studies examining molecular diagnostics, resistance gene profiling, and surveillance methods have increased as a result.

Antimicrobial peptides (AMPs) and phage therapy have also become novel substitutes for traditional antibiotics, providing pathogen-specific and biofilm-disruptive effects that may help treat MDR infections (Serrano et al., 2023; Ramirez-Acuña et al., 2019).

Quorum sensing inhibitors and quorum quenching enzymes are examples of other frontier areas that seek to interfere with bacterial communication pathways that are necessary for biofilm maturation and virulence. These new subdomains demonstrate a paradigm shift toward focused, interdisciplinary strategies meant to address the intricate pathophysiology of *P. aeruginosa* in diabetic foot infections and lessen the threat of antibiotic resistance worldwide.

### 6.3 Global distribution of *P. aeruginosa* in diabetic foot infection research and the patterns of collaboration

Significant differences in research productivity and collaboration can be seen in the global distribution of studies on *Pseudomonas aeruginosa* in diabetic foot infections (DFIs). Most publications come from high-output nations like China, India, and the United States, which together control the majority of research worldwide because of their high rates of disease, strong funding systems, advanced diagnostic capabilities, and sophisticated research infrastructure (Saeedi et al., 2020; Aminabee et al., 2024). Structural disparities in research participation worldwide are highlighted by the underrepresentation of regions like sub-Saharan Africa, where Egypt is the only significant contributor (Saqib & Rafique, 2021). Studies from regions where the clinical burden of *P. aeruginosa* is probably high but research capacity is low are underrepresented, as the distribution of research is further distorted by institutional and technological capabilities (Saeedi et al., 2020).

Additionally, bibliometric analyses reveal that the majority of research is still country-specific, as evidenced by the high number of Single Country Publications (SCPs) relative to Multi-Country Publications (MCPs), which suggests little international cooperation. In contrast to the United States, which reported 32 SCPs and 4 MCPs, India reported 42 SCPs with only 5 MCPs. According to the collaboration network visualization, Saudi Arabia and Canada, despite producing fewer publications, have stronger collaborative ties than the United States and China, which lead in publication volume but have relatively weak cross-national ties. Additionally, the co-authorship network demonstrates that although international partnerships are dominated by five major clusters, many countries—particularly in the Middle East and Africa—remain isolated or only loosely connected (Zhu & Liu, 2020; Gazni et al., 2012). In order to advance equity and optimize worldwide innovation in the fight against antibiotic resistance and diabetic foot complications, these findings highlight the necessity of bolstering transnational research collaborations and capacity building, especially in low- and middle-income nations.

### 6.4 Research Frontiers and Thematic Evolution

In response to new clinical requirements, advancements in technology, and global health priorities, the scientific landscape of Pseudomonas aeruginosa research in diabetic foot infections (DFIs) has changed into a number of discrete thematic clusters, each of which has developed chronologically. Molecular Mechanisms, Diagnostics, Treatment Strategies, and Epidemiology are the four main clusters identified by the co-occurrence and thematic evolution analyses. The following describes each cluster using insights from temporal overlay visualization and keyword burst patterns.

#### 1. Molecular Mechanisms

With a particular emphasis on biofilm formation, virulence factors, and antibiotic resistance mechanisms, this cluster investigates the genetic, biochemical, and physiological underpinnings of *P. aeruginosa* pathogenicity.

**1985–2013**: The main topics of early research were biofilm formation and general microbial virulence (e.g., pyocyanin, proteases, rhamnolipids). Antibacterial activity, bacterium isolation, and Staphylococcus aureus were important terms.

**2014-2021**: Attention turned to quorum sensing systems (Las, Rhl, Pqs, and Iqs), which control biofilm formation and virulence. Research on the mechanisms underlying antibiotic resistance also increased during this time, particularly with regard to multidrug resistance (MDR) and extended drug-resistant (XDR) phenotypes caused by beta-lactamase production, efflux pumps, and porin loss.

**2020–2024**: "Burst keywords" like quorum sensing, quorum quenching, and metagenomics are becoming more common, and genomics-based concepts are emerging quickly. The investigation of genetic control and manipulation was broadened by the CRISPR mechanism and regulatory small RNA signal.

#### 2. Diagnostics

The need for quicker and more precise identification of resistant strains in chronic wounds has fueled advancements in diagnostics.

**1985-2013**: Basic culture methods predominated, with sporadic mentions of polymicrobial interactions and wound microbiology.

**2014–2021**: Rapid molecular diagnostics, including PCR-based assays for resistance determinants, saw an increase in citation bursts. Keywords like bacteriology, microbial spectrum, and in vitro susceptibility analysis gained prominence.

**2022-2024**: Overlay analysis reveals the use of big data-driven methods to track *P. aeruginosa* outbreaks in diabetic foot infection cohorts and the emergence of next-generation sequencing for epidemiological tracing, which provides higher-resolution insights into pathogen diversity and resistance. A popular term that reflects a movement toward comprehending intricate microbial communities is metagenomics.

#### 3. Treatment Strategies

In order to combat MDR *P. aeruginosa*, treatment research has shifted from empirical therapy to mechanism-based, focused interventions.

**1985-2013**: There was a thematic emphasis on conventional antibiotics, with a limited emphasis on resistance profiles and a focus on the empirical use of broad-spectrum antibiotics (such as vancomycin, ciprofloxacin, and amikacin). Additionally, amputation results and wound care procedures were taken into account.

**014–2021:** The emergence of extensively drug-resistant (XDR) and multidrug-resistant (MDR) strains sparked interest in biofilm-disrupting agents, combination therapy, silver nanoparticles, and novel antimicrobials.

**2022–2024:** Keywords such as phage therapy, silver nanoparticles, antimicrobial peptides (AMPs), and quorum quenching enzymes indicate the trend toward novel, unconventional treatments. As a result of the interest in bioengineered wound dressings, nisin Z, chitosan composites, and hydrogels were also noted as popular terms during this time. Frontier concepts are also beginning to emerge in adjunctive therapies that use immune modulation and CRISPR-based gene editing.

#### 4. Epidemiology and Public Health

Prevalence trends, risk factors, and health system difficulties in treating P. aeruginosa infections are covered in this cluster.

**1985-2013**: Descriptive research on the prevalence and mortality of diabetic foot complications was conducted.

**2014-2021**: Research started looking at hospital-based outbreaks, comorbidities, and geographic patterns. Terms such as MRSA, DFU, osteomyelitis, and polymicrobial infection are more central in thematic maps.

**2022–2024:** A strategic shift toward prevention, policy-making, and research integration is reflected in buzzwords like antimicrobial stewardship, AMR surveillance, and global collaboration networks. Disparities in research participation, particularly from low- and middle-income nations, are highlighted. A greater emphasis on the effects of health systems, social determinants, and the interaction of co-infecting species is indicated by thematic evolution.

### 6.5 Newly Emerging Concepts (“Burst Keywords”)

A number of recently developed ideas and technologies that are influencing the field are highlighted by keyword burst analysis and bibliometric overlay visualization:

#### Quorum quenching

Interfering with bacterial communication to stop the growth of biofilms and the spread of infection.

#### CRISPR

Genome editing technologies for quick diagnostics and pathogen manipulation.

#### Metagenomics

Profiling the microbial communities in polymicrobial diabetic foot infections at high throughput.

#### Antimicrobial peptides

Alternative treatments to deal with MDR strains.

#### Phage therapy

the use of *P. aeruginosa*-specific bacteriophages as an adjuvant or substitute for antibiotics.

These outbursts demonstrate the continuous thematic development, signifying a move away from descriptive and traditional clinical research and toward creative, interdisciplinary methods intended to tackle challenging clinical issues.

## CONCLUSION

This thorough systematic review and bibliometric analysis demonstrates how *Pseudomonas aeruginosa* in diabetic foot infections has significantly changed and intensified worldwide, especially in the past two decades. The field is now characterized by rapid growth and thematic specialization, particularly in biofilm research, antimicrobial resistance, and emerging therapies like phage therapy and quorum quenching, from sporadic, broad-based studies. Notwithstanding these developments, the distribution of research output is still uneven, with low-resource regions continuing to be underrepresented and the majority of contributions coming from a small number of nations with robust research infrastructure. The need for more strong, inclusive international partnerships is highlighted by the mapping of global collaboration patterns, which shows a preponderance of intra-country efforts and fragmented inter-regional interactions. When taken as a whole, these results highlight both the advancements made and the ongoing deficiencies in cooperation and knowledge creation. To address the persistent problems caused by *P. aeruginosa* in diabetic foot infections and enhance patient outcomes globally, future research should strive to promote greater research equity and holistic, cross-disciplinary innovation.

## Data Availability

All data produced in the present study are available upon request to the authors

## References

Abdullah, M., Humayun, A., Imran, M., Bashir, M. A., & Malik, A. A. (2022). A bibliometric analysis of global research performance on tuberculosis (2011–2020): Time for a global approach to support high-burden countries. Journal of Family and Community Medicine, 29(2), 117–124.

Afrifa, S., Zhang, T., Appiahene, P., & Varadarajan, V. (2022). Mathematical and machine learning models for groundwater level changes: a systematic review and bibliographic analysis. Future Internet, 14(9), 259.

Al-Joufi, F. A., Aljarallah, K. M., Hagras, S. A., Al Hosiny, I. M., Salem-Bekhit, M. M., Youssof, A. M. E., & Shakeel, F. (2020). Microbial spectrum, antibiotic susceptibility profile, and biofilm formation of diabetic foot infections (2014–18): a retrospective multicenter analysis. 3 Biotech, 10(7). 10.1007/s13205-020-02318-x

Alkhudhairy, M. K., & Al-Shammari, M. M. M. (2020). Prevalence of metallo-β-lactamase– producing Pseudomonas aeruginosa isolated from diabetic foot infections in Iraq. New microbes and new infections, 35, 100661.

Álvarez-García, J., Maldonado-Erazo, C. P., Río-Rama, D., Cruz, M., and Castellano-Álvarez, F. J. (2019). Cultural Heritage and Tourism Basis for Regional Development: Mapping of Scientific Coverage. Sustainability, 11(21), 6034. 10.3390/su11216034

Aminabee, S., Karnati, S., Shankar, K. R., Devi, T. S., Srinivasu, P., & Harshini, G. (2024). Exploring Diabetes Research Trends in India: A Comprehensive Bibliometric Analysis Using VOSviewer. Biomedical and Pharmacology Journal, 17(3), 1655–1666.

Aria, M., & Cuccurullo, C. (2017). bibliometrix: An R-tool for comprehensive science mapping analysis. Journal of informetrics, 11(4), 959–975.

Baker, H. K., Kumar, S., & Pandey, N. (2021). Forty years of the Journal of Futures Markets: A bibliometric overview. Journal of Futures Markets, 41(7), 1027–1054.

Bornmann, L., Wagner, C., & Leydesdorff, L. (2015). BRICS countries and scientific excellence: A bibliometric analysis of most frequently cited papers. Journal of the Association for Information Science and Technology, 66(7), 1507–1513. 10.1002/asi.23333

Boulton, A. J., Vileikyte, L., Ragnarson-Tennvall, G., & Apelqvist, J. (2005). The global burden of diabetic foot disease. The Lancet, 366(9498), 1719–1724.

Chichón, G., López, M., de Toro, M., Ruiz-Roldán, L., Rojo-Bezares, B., & Sáenz, Y. (2023). Spread of Pseudomonas aeruginosa ST274 clone in different niches: resistome, virulome, and phylogenetic relationship. Antibiotics, 12(11), 1561.

Citron, D. M., Goldstein, E. J., Merriam, C. V., Lipsky, B. A., & Abramson, M. A. (2007). Bacteriology of moderate-to-severe diabetic foot infections and in vitro activity of antimicrobial agents. Journal of clinical Microbiology, 45(9), 2819–2828.

Cobo, M. J., López-Herrera, A. G., Herrera-Viedma, E., & Herrera, F. (2011). Science mapping software tools: Review, analysis, and cooperative study among tools. Journal of the American Society for information Science and Technology, 62(7), 1382–1402.

De Oliveira, D. M., Forde, B. M., Kidd, T. J., Harris, P. N., Schembri, M. A., Beatson, S. A., … & Walker, M. J. (2020). Antimicrobial resistance in ESKAPE pathogens. Clinical microbiology reviews, 33(3), 10–1128.

Deng, Pin, Hongshuo Shi, Xuyue Pan, Huan Liang, Shulong Wang, Junde Wu, Wei Zhang et al. "Worldwide research trends on diabetic foot ulcers (2004–2020): suggestions for researchers." Journal of Diabetes Research 2022, no. 1 (2022): 7991031.

Domingues, M. M., Felício, M. R., & Gonçalves, S. (2019). Antimicrobial peptides: effect on bacterial cells. Atomic Force Microscopy: Methods and Protocols, 233–242.

Fuentes-Peñaranda, Y., Labarta-González-Vallarino, A., Arroyo-Bello, E., & Gómez de Quero Córdoba, M. (2025). Global Trends in Diabetic Foot Research (2004–2023): A Bibliometric Study Based on the Scopus Database. International Journal of Environmental Research and Public Health, 22(4), 463.

Garousi, M., MonazamiTabar, S., Mirazi, H., Farrokhi, Z., Khaledi, A., & Shakerimoghaddam, A. (2023). Epidemiology of Pseudomonas aeruginosa in diabetic foot infections: a global systematic review and meta-analysis. Germs, 13(4), 362.

Gazni, A., Sugimoto, C. R., & Didegah, F. (2012). Mapping world scientific collaboration: Authors, institutions, and countries. Journal of the American Society for Information Science and Technology, 63(2), 323–335. 10.1002/asi.21688

Gonzales-Malca, J. A., Tirado-Kulieva, V. A., Abanto-López, M. S., Aldana-Juárez, W. L., & Palacios-Zapata, C. M. (2022). Bibliometric analysis of research on the main genes involved in meat tenderness. Animals, 12(21), 2976.

Hallinger, P., Gümüş, S., & Bellibaş, M. Ş. (2020). ’Are principals instructional leaders yet?’A science map of the knowledge base on instructional leadership, 1940–2018. Scientometrics, 122(3), 1629–1650.

Hamid, S. F., Taha, A. B., & Abdulwahid, M. J. (2020). Distribution of blaTEM, blaSHV, blaCTX-M, blaOXA, and blaDHA in Proteus mirabilis isolated from diabetic foot infections in Erbil, Iraq. Cellular and Molecular Biology, 66(1), 88–94.

Lee, J. H., Kim, N. H., Jang, K. M., Jin, H., Shin, K., Jeong, B. C., … & Lee, S. H. (2023). Prioritization of critical factors for surveillance of the dissemination of antibiotic resistance in Pseudomonas aeruginosa: A systematic review. International Journal of Molecular Sciences, 24(20), 15209.

Lipsky, B. A., Berendt, A. R., Cornia, P. B., Pile, J. C., Peters, E. J., Armstrong, D. G., … & Senneville, E. (2012). 2012 Infectious Diseases Society of America clinical practice guideline for the diagnosis and treatment of diabetic foot infections. Clinical infectious diseases, 54(12), e132–e173.

Lipsky, B. A., Senneville, É., Abbas, Z. G., Aragón-Sánchez, J., Diggle, M., Embil, J. M., … & International Working Group on the Diabetic Foot (IWGDF). (2020). Guidelines on the diagnosis and treatment of foot infection in persons with diabetes (IWGDF 2019 update). Diabetes/metabolism research and reviews, 36, e3280.

Macdonald, K. E., Boeckh, S., Stacey, H. J., & Jones, J. D. (2021). The microbiology of diabetic foot infections: A global meta-analysis. BMC Infectious Diseases, 21, Article 770. 10.1186/s12879-021-06516-7

Maflahi, N. and Thelwall, M. (2015). When are readership counts as useful as citation counts? Scopus versus Mendeley for LIS journals. Journal of the Association for Information Science and Technology, 67(1), 191–199. 10.1002/asi.23369

Maity, S., Leton, N., Nayak, N., Jha, A., Anand, N., Thompson, K., … & Nauhria, S. (2024). A systematic review of diabetic foot infections: pathogenesis, diagnosis, and management strategies. Frontiers in Clinical Diabetes and Healthcare, 5, 1393309.

Moher, D., Liberati, A., Tetzlaff, J., & Altman, D. G. (2009). Preferred reporting items for systematic reviews and meta-analyses: the PRISMA statement. Bmj, 339.

Mojica, M. F., Rossi, M. A., Vila, A. J., & Bonomo, R. A. (2022). The urgent need for metallo-β-lactamase inhibitors: an unattended global threat. The Lancet Infectious Diseases, 22(1), e28–e34. 10.1016/S1473-3099(20)30868-9

Moral-Muñoz, J. A., Herrera-Viedma, E., Santisteban-Espejo, A., & Cobo, M. J. (2020). Software tools for conducting bibliometric analysis in science: An up-to-date review. Profesional de la Información, 29(1).

Ramirez-Acuña, J. M., Cardenas-Cadena, S. A., Marquez-Salas, P. A., Garza-Veloz, I., Perez-Favila, A., Cid-Baez, M. A., Flores-Morales, V., & Martinez-Fierro, M. L. (2019). Diabetic Foot Ulcers: Current Advances in Antimicrobial Therapies and Emerging Treatments. Antibiotics, 8(4), 193. 10.3390/antibiotics8040193

Rejeb, A., Rejeb, K., Abdollahi, A., Zailani, S., Iranmanesh, M., and Ghobakhloo, M. (2021). Digitalization in food supply chains: a bibliometric review and key-route main path analysis. Sustainability, 14:83. doi: 10.3390/su14010083

Resurchify. (2024). Journal H-index rankings. Retrieved from https://www.resurchify.com/

SCImago Journal Rank. (2024). Journal rankings and metrics. Retrieved from https://www.scimagojr.com/

Rhoads, D. D., Wolcott, R. D., Sun, Y., Dowd, S. E. (2013). Biofilms in diabetic foot ulcers: Significance and clinical relevance. Microorganisms, 8(10), 1580. 10.3390/microorganisms8101580

Rojas-Sánchez, M. A., Palos-Sánchez, P. R., & Folgado-Fernández, J. A. (2023). Systematic literature review and bibliometric analysis on virtual reality and education. Education and Information Technologies, 28(1), 155–192.

Saeedi, P., Petersohn, I., Salpea, P., Malanda, B., Karuranga, S., Unwin, N., & IDF Diabetes Atlas Committee. (2019). Global and regional diabetes prevalence estimates for 2019 and projections for 2030 and 2045: Results from the International Diabetes Federation Diabetes Atlas. Diabetes research and clinical practice, 157, 107843.

Sannathimmappa, M. B., Nambiar, V., Aravindakshan, R., Al Khabori, M. S. J., Al-Flaiti, A. H. S., Al-Azri, K. N. M., … & Al Kiyumi, A. R. M. (2021). Diabetic foot infections: Profile and antibiotic susceptibility patterns of bacterial isolates in a tertiary care hospital of Oman. Journal of Education and Health Promotion, 10(1), 254

Saqib, M. A. N., & Rafique, I. (2021). Health research funding and its output in Pakistan. Eastern Mediterranean Health Journal, 27(9), 906–910. 10.26719/emhj.21.038

Serra, R., Grande, R., Butrico, L., Rossi, A., Settimio, U. F., Caroleo, B., & De Franciscis, S. (2015). Chronic wound infections: the role of Pseudomonas aeruginosa and Staphylococcus aureus. Expert review of anti-infective therapy, 13(5), 605–613.

Serrano, I., Alhinho, B., Cunha, E., Tavares, L., Trindade, A., & Oliveira, M. (2023). Bacteriostatic and antibiofilm efficacy of a Nisin Z solution against co-cultures of Staphylococcus aureus and Pseudomonas aeruginosa from diabetic foot infections. Life, 13(2), 504.

Sganzerla, W. G., Ampese, L. C., Mussatto, S. I., & Forster-Carneiro, T. (2021). A bibliometric analysis on potential uses of brewer’s spent grains in a biorefinery for the circular economy transition of the beer industry. Biofuels, Bioproducts and Biorefining, 15(6), 1965–1988.

Silue, Y., & Fawole, O. A. (2024). Global research network analysis of edible coatings and films for preserving perishable fruit crops: current status and future directions. Foods, 13(15), 2321.

Sweileh, W. M., Al-Jabi, S. W., Zyoud, S. H., Sawalha, A. F., & Abu-Taha, A. S. (2018). Global research output in antimicrobial resistance among uropathogens: A bibliometric analysis (2002– 2016). Journal of Global Antimicrobial Resistance, 13, 104–114. 10.1016/j.jgar.2017.11.017

Van Eck, N., & Waltman, L. (2010). Software survey: VOSviewer, a computer program for bibliometric mapping. scientometrics, 84(2), 523–538.

Wolcott, R. D., Hanson, J. D., Rees, E. J., Koenig, L. D., Phillips, C. D., Wolcott, R. A., … & White, J. S. (2016). Analysis of the chronic wound microbiota of 2,963 patients by 16S rDNA pyrosequencing. Wound repair and regeneration, 24(1), 163–174.

World Health Organization (WHO). (2021). Global antimicrobial resistance and use surveillance system (GLASS) report 2021. https://www.who.int/publications/i/item/9789240027336

Zhao, X. P., Li, D., Li, C. L., Zhang, Y. N., Zhao, N. R., & Xu, J. X. (2023). Knowledge mapping of diabetic foot research based on Web of Science database: a bibliometric analysis. Medicine, 102(26), e34053.

Zhu, J., & Liu, W. (2020). *Comparing like with like: China ranks first in SCI-indexed research articles since* 2018. Scientometrics, 124(2), 1691–1700. 10.1007/s11192-020-03525-2

